# Multi-site validation of a functional assay to adjudicate *SCN5A* Brugada Syndrome-associated variants

**DOI:** 10.1101/2023.12.19.23299592

**Authors:** Joanne G. Ma, Matthew J. O’Neill, Ebony Richardson, Kate L. Thomson, Jodie Ingles, Ayesha Muhammad, Joseph F. Solus, Giovanni Davogustto, Katherine C. Anderson, M. Benjamin Shoemaker, Andrew B. Stergachis, Brendan J. Floyd, Kyla Dunn, Victoria N. Parikh, Henry Chubb, Mark J. Perrin, Dan M. Roden, Jamie I. Vandenberg, Chai-Ann Ng, Andrew M. Glazer

## Abstract

Brugada Syndrome (BrS) is an inheritable arrhythmia condition that is associated with rare, loss-of-function variants in the cardiac sodium channel gene, *SCN5A*. Interpreting the pathogenicity of *SCN5A* missense variants is challenging and ∼79% of *SCN5A* missense variants in ClinVar are currently classified as Variants of Uncertain Significance (VUS). An *in vitro SCN5A*-BrS automated patch clamp assay was generated for high-throughput functional studies of Na_V_1.5. The assay was independently studied at two separate research sites – Vanderbilt University Medical Center and Victor Chang Cardiac Research Institute – revealing strong correlations, including peak *I*_Na_ density (*R*^2^=0.86). The assay was calibrated according to ClinGen Sequence Variant Interpretation recommendations using high-confidence variant controls (n=49). Normal and abnormal ranges of function were established based on the distribution of benign variant assay results. The assay accurately distinguished benign controls (24/25) from pathogenic controls (23/24). Odds of Pathogenicity values derived from the experimental results yielded 0.042 for normal function (BS3 criterion) and 24.0 for abnormal function (PS3 criterion), resulting in up to strong evidence for both ACMG criteria. The calibrated assay was then used to study *SCN5A* VUS observed in four families with BrS and other arrhythmia phenotypes associated with *SCN5A* loss-of-function. The assay revealed loss-of-function for three of four variants, enabling reclassification to likely pathogenic. This validated APC assay provides clinical-grade functional evidence for the reclassification of current VUS and will aid future *SCN5A*-BrS variant classification.

## Introduction

Brugada Syndrome (BrS) is an autosomal dominant disease characterized by ST-segment elevation in the right precordial leads (V1-V3) of the electrocardiogram (ECG).^1^ Patients with the definitive (“Type 1”) BrS ECG pattern are at increased risk of sudden cardiac death due to ventricular fibrillation.^2^ Sudden cardiac death may be the first disease manifestation, making early identification of at-risk patients critical. Up to 30% of BrS patients have rare variants in *SCN5A*, which encodes Na_V_1.5, the cardiac voltage-gated sodium channel.^3^ BrS-associated *SCN5A* variants are loss-of-function (LOF) and decrease cardiac excitability and reduce electrical conduction velocity. A ClinGen expert panel asserted that *SCN5A* is the only gene for which rare variants are definitively associated with BrS.^4^ BrS is also substantially influenced by common genetic variation, with 12 common loci identified throughout the genome in a recent genome-wide association study.^5^

Genetic testing offers the ability to identify individuals at risk for BrS, and proactively optimize medical management before they experience life-threatening cardiac events.^6,7^ However, classification of variants in *SCN5A* remains challenging as *SCN5A* variants can have normal function or cause LOF or gain-of-function (GOF). All variants are currently classified by the ACMG/AMP v3 framework as benign (B), likely benign (LB), variant of uncertain significance (VUS), likely pathogenic (LP), or pathogenic (P).^8^ Unfortunately, ∼79% of missense *SCN5A* variants in ClinVar are currently classified as VUS,^9,10^ This high burden of VUS has resulted in underdiagnosis, misdiagnosis, or inappropriate interventions.^11,12^

High-throughput functional assays provide an opportunity to improve variant classifications.^8^ Manual patch clamping is the gold-standard method for measuring the function of ion channel variants. However, patch clamping is low-throughput, and individual reports usually describe functional characteristics for one or a small number of variants compared to wild-type, precluding assay calibration and generalization across laboratories. High-throughput automated patch-clamp (APC) electrophysiology platforms enable the rapid functional assessment of large numbers of ion channel variants.^13–17^ In the 2015 v3 ACMG/AMP guidelines, well-established functional assay results were applied at a strong level.^8^ These criteria are termed BS3 for a normal assay result or PS3 for an abnormal assay result. In 2020, the ClinGen Sequence Variant Interpretation (SVI) Working Group recommended instead applying the ACMG/AMP functional assay criteria at a range of evidence strengths depending on the assay’s performance on pathogenic and benign variant controls.^18^ Jiang et al.^16^ previously deployed this framework on a APC assay for *KCNH2*-related Long QT Syndrome, with functional data initially providing moderate-strength evidence for clinical classifications.^16^ This *KCNH2* assay was recently upgraded to strong evidence strength by including more variant controls.^19^ This framework has not yet been applied to any other APC assays, such as an APC assay for Brugada Syndrome (*SCN5A*-BrS). Furthermore, multi-site replication of APC assay calibration has not yet been reported, an important step in promoting reproducibility.

In this study, we calibrate an *SCN5A*-BrS APC assay using a large set of P/LP and B variant controls. We demonstrate high replicability of this assay between two independent research centers and high concordance with clinical classifications. We then used the calibrated assay to provide functional evidence for reclassification of *SCN5A* VUS identified in clinical cases of BrS and other severe arrhythmias.

## Materials and Methods

### *SCN5A* transcript

Variants were annotated and experimentally studied using the MANE Select *SCN5A* transcript ENST00000423572.7 (NM_000335.5), the most common transcript in the adult heart.^20^ This 2015-amino acid transcript includes the adult exon 6A and does not include the alternatively spliced Gln1077 residue. In some previous studies, variants have been annotated according to the 2016-amino acid transcript that includes adult exon 6A and contains Gln1077, ENST00000333535.^17,21^ Table S1 shows variants annotated according to both reference transcripts.

### Classification of *SCN5A* variants

Variant controls were classified using the 2015 ACMG/AMP v3 criteria for variant interpretation^8^ by a team of clinical geneticists and cardiac genetic counselors. Candidate *SCN5A* variants were selected from the ClinVar^9^ and gnomAD variant databases (v2.1.1 and v3.1.2).^22^ For the classification of benign variants, the maximum credible allele frequency of *SCN5A* variants in population databases was defined using the allele frequency app (http://cardiodb.org/allelefrequencyapp/)^23^ with an inputted BrS prevalence of 1 in 2000,^24^ allele heterogeneity of 0.02, genetic heterogeneity of 0.3, and penetrance set to the lowest setting of 0.01. Using these thresholds, BA1 can be applied as the stand-alone criteria for all variants meeting a maximum credible population allele frequency of 0.00015.^25^ However, for identifying control benign variants, we conservatively applied BA1 to variants with a maximum credible allele frequency of 0.0003 in gnomAD. P/LP variants for BrS were classified with multiple ACMG/AMP criteria,^26^ including the number of probands with consistent disease phenotype (PS4), absence or rarity of allele in large scale population databases (maximum allele frequency of 3 in 100,000 alleles; PM2_supporting), location of variant (PM1),^27^ *in silico* predictors (PP3) including Metascores (MetaLR, MetaSVM, MetaRNN, REVEL^28^) and VarSome^29^, and segregation data (PP1). Clinical databases from the Centre for Population Genomics and Oxford Genetics Laboratories provided additional proband evidence for pathogenic classifications. The final set of 25 B variant controls is shown in Table S2 and the final set of 24 P/LP variant controls is shown in Table S3.

### *SCN5A* automated patch clamp assay

We followed previously reported protocols to independently generate stable HEK293 variant cell lines (Victor Chang Cardiac Research Institute, VCCRI;^16,30^ Vanderbilt University Medical Center, VUMC).^17,31^ Briefly, *SCN5A* variant constructs were cloned or subcloned independently using a QuikChange Lightning Mutagenesis kit (Agilent) into a AttB:SCN5A:IRES:mCherry-blasticidinR plasmid (VUMC), or into pcDNA^TM^5/FRT/TO (Thermo Fisher Scientific) by Genscript Inc. (Piscataway, USA) (VCCRI). See Table S4 for a description of all primers used in this study. Plasmids encoding different variants were transfected into modified HEK293 cell lines at each site. VUMC used a “negative selection” landing pad system (a gift from Kenneth Matreyek and Doug Fowler).^32^ HEK293 negative selection landing pad cells were co-transfected with a AttB-containing *SCN5A* variant plasmid and a plasmid bearing Bxb1 recombinase (a gift from Pawel Pelczar^33^, Addgene plasmid #51271) with Lipofectamine 2000 (Thermo Fisher Scientific) following manufacturer’s instructions. Cells were grown in selection media containing 5 ug/mL doxycycline (to induce promoter expression; Sigma-Aldrich), 100 ug/mL blasticidin S (to eliminate cells not expressing the blasticidin-resistant plasmid; Sigma-Aldrich), and 10 nM AP1903 (to eliminate non-integrated cells expressing a AP1903-inducible caspase gene; MedChemExpress). Cells were grown for 10 days and screened by flow cytometry to confirm expression levels. VCCRI generated stable cell lines by co-transfecting an *SCN5A* variant-expressing pcDNA^TM^5/FRT/TO plasmid with a pOG44 Flp-Recombinase Expression Vector (Thermo Fisher Scientific). The plasmids were transfected with Lipofectamine 3000 (Thermo Fisher Scientific) following manufacturer’s instructions. Cell lines were grown in selection media containing 200 ug/mL of Hygromycin (Thermo Fisher Scientific) and 10 ug/mL Blasticidin (InvivoGen) for up to 14 days, and *SCN5A* expression was induced with 200 ng/mL doxycycline (Sigma-Aldrich) 48 hours prior to experimentation. Electrophysiology experiments at both sites were conducted on the SyncroPatch 384 PE (Nanion Technologies, Munich, Germany) using Nanion chips containing *SCN5A* wild-type (WT) and variant control cell lines. Data analysis was performed using site-specific, custom *SCN5A* analysis scripts in R (VUMC) and Matlab (VCCRI). See Figure 1 for an overview of the methods used in this project. See Supplemental Methods and Figure S1 for detailed electrophysiology protocols and site-specific methods.

**Figure 1:**
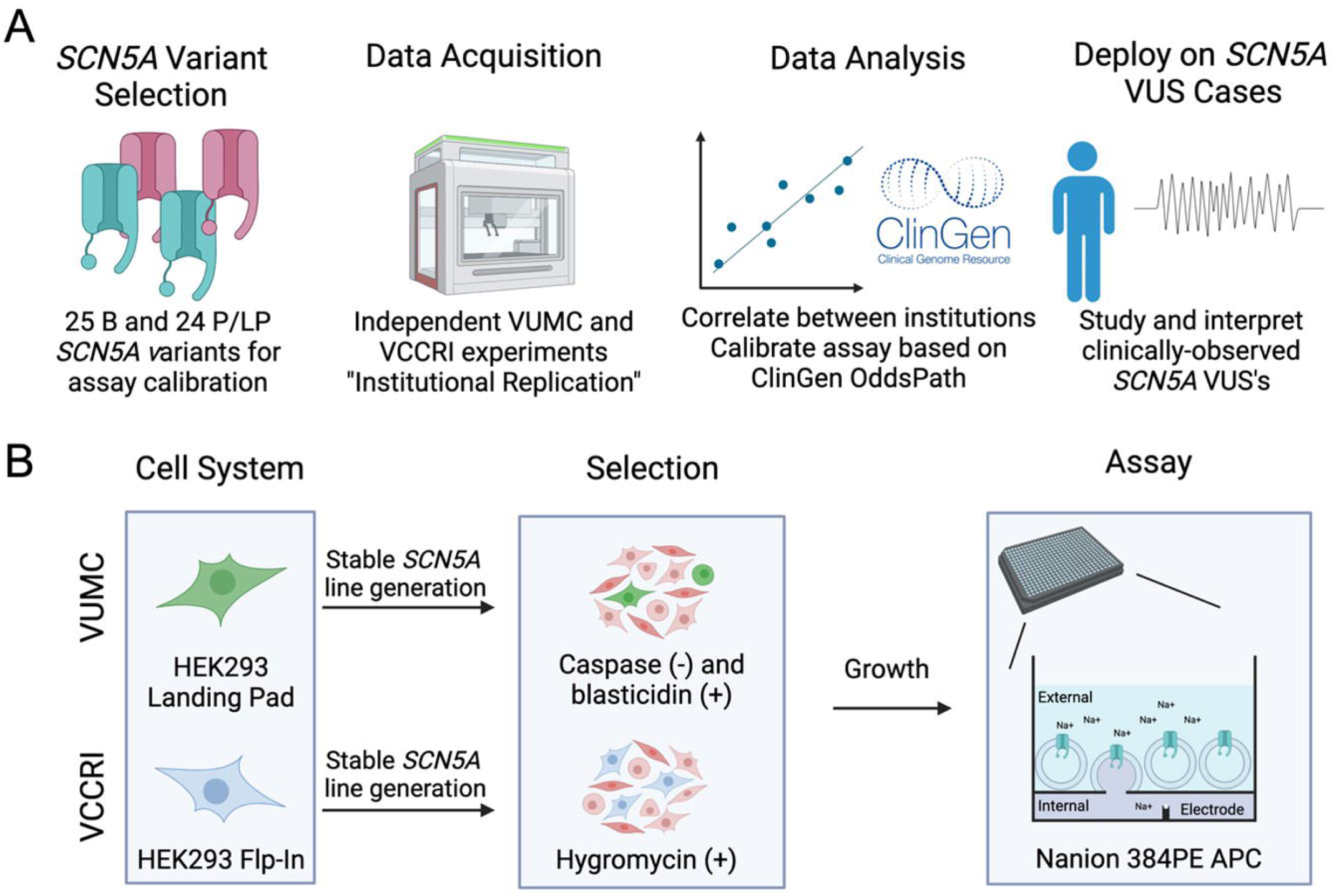
A Calibrated *SCN5A* Automated Patch Clamp Assay. (A) Overview of project design. Selection of calibration variants, independent execution of experiments at two sites, calibration of results, and application to clinical cases. (B) Individual site assay schematic. Minor differences in cell systems, selection drugs, and APC conditions/analysis existed between sites and are described in the Methods.

### Quality control, sample size, and reproducibility

Both sites implemented common quality control criteria for seal resistance (R_seal_), capacitance (C_slow_), series resistance (R_series_) and residual background current after leak correction. We also performed QC for time to peak as a surrogate for adequate voltage-clamp control. Lastly, current amplitude thresholds were applied during analysis of channel gating parameters: steady state activation (SSA), steady state inactivation (SSI), and recovery from inactivation (RFI). Specific QC parameters applied at each site are presented in Supplemental Methods.

Power calculations for sample sizes were performed based on cells expressing WT *SCN5A* that passed QC controls. Sample size calculation was performed (see Supplemental Methods) to detect a 25% difference at 90% power, where LJ = 0.05. Tests of normality were implemented with Shapiro-Wilk tests. Non-normal variables were transformed with a square root-transformation, then normalized to the WT mean or individual chips or groups of chips to control for plate-to-plate and batch-to-batch variation, respectively.

### Z-score and OddsPath calculations

For each functional property, normalized means of WT and benign variants were combined to establish thresholds to distinguish variants with normal function from variants with abnormal function. Z-scores were calculated (Equation 1) for all measured values to quantify each variant’s function, with smaller Z-scores associated with lower values (such as lower current density).

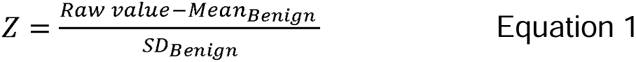

For *I*_Na_ density, variants with Z-scores between –2 and +2 were considered to have normal function. For *I*_Na_ density, variants with a Z-score between –2 and –4 were considered moderate LOF and variants with a Z-score below –4 were considered severe LOF ^19^. For SSA, SSI, and RFI, variants with Z > 4 or Z < –4 were considered abnormal. The evidence strength for each *SCN5A* functional property was calculated according to the Odds of Pathogenicity (OddsPath) equations as recommended by the ClinGen Sequence Variant Interpretation working group ^18,26^. A spreadsheet detailing the OddsPath calculations is presented in Table S8.

### Analysis of channel functional properties

*SCN5A* functional properties analyzed in this assay were peak sodium current (*I*_Na_) density, SSA, SSI and RFI. Diagrams of voltage protocols are presented in Figure S1 and equations for analysis used in this study are presented in the Supplementary Methods. The *I*_Na_ density was measured at –30 mV following a holding potential of either –120 mV (VUMC and VCCRI) or –90 mV (VCCRI). After quality control, activating current amplitude was divided by cell capacitance to calculate current density normalized to cell size, measured in pA/pF. SSA was obtained by measuring currents in cells depolarized from a holding potential of –120 mV to voltages in the range –100 mV to 0 mV, at 5 mV increments. SSA was fit to the Boltzmann equation using nonlinear least-squares to determine the voltage at which half of the channels were activated (SSA V_50_). SSI was determined after holding channels at voltages in the range –140 mV to –40 mV for 500 ms (in 5 mV increments) then measuring tail current amplitudes recorded at –30 mV. These values were fit to the Boltzmann equation to determine SSI V_50_. RFI was determined by the ratio of current amplitude at a test pulse to current amplitude at a pre-pulse with increasing time intervals between the two pulses. The time at which channels were half recovered (T_50_) was used for analysis.

### Clinical evaluation and genetic testing

*SCN5A* VUS were identified in patients with BrS diagnoses or severe cardiac conduction disease from four tertiary care centers: Vanderbilt University Medical Center, Stanford Medicine, University of Washington Hospital, and The Royal Melbourne Hospital. All studies were approved by local Institutional Review Boards/Research Ethics Committees and informed consent was obtained as required by these entities. Clinical information was obtained through retrospective review of medical records. Patients underwent commercial genetic testing using gene-panels. Functional characterization of each clinical *SCN5A* VUS using the calibrated assay was performed as described above. Although the APC OddsPath calculations supported the use of BS3_strong, we conservatively applied a graded evidence strength scheme. Z-scores of the square root transformed current density values were used to calculate the appropriate ACMG criterion (PS3_strong for Z < –4, PS3_moderate for –4 < Z < –3, PS3_supporting for –3 < Z < –2, BS3_supporting for –2 < Z < –1, and BS3_moderate for Z > –1). We note that we conservatively applied BS3_moderate evidence strength for variants with –1 < Z < 1 and BS3_supporting for –2 < Z < –1 and Z > 1. This was due to caution over misclassifying a disease-causing variant as benign or likely benign, and because we have not yet calibrated the assay to detect other types of *SCN5A* dysfunction such as GOF. Variant reclassification was independently performed at each clinical site by a genetic counselor, clinical geneticist, or physician specializing in genomic medicine.

## Results

### Square root transformation, power calculation

We measured typical *I*_Na_ currents from 2,727 cells expressing WT *SCN5A* (Figure 2A-B). The raw current densities showed a non-Gaussian distribution (Figure 2C-D, p<0.0001, Shapiro-Wilk test). The current density values were converted to a more normal distribution by application of a square-root transformation (Figure 2E-F; p=0.304 and p<0.0001 at VCCRI and VUMC, respectively; Shapiro-Wilk test).^16,34^ Using a sample size power calculation (see Supplemental Equations), we determined that a minimum of 30 cells per variant was required to detect a 25% difference in current density at 90% power with a 95% confidence interval. We therefore measured at least 30 cells per site for each variant with replicates spanning independent transfections, chips, and experimental days.

**Figure 2:**
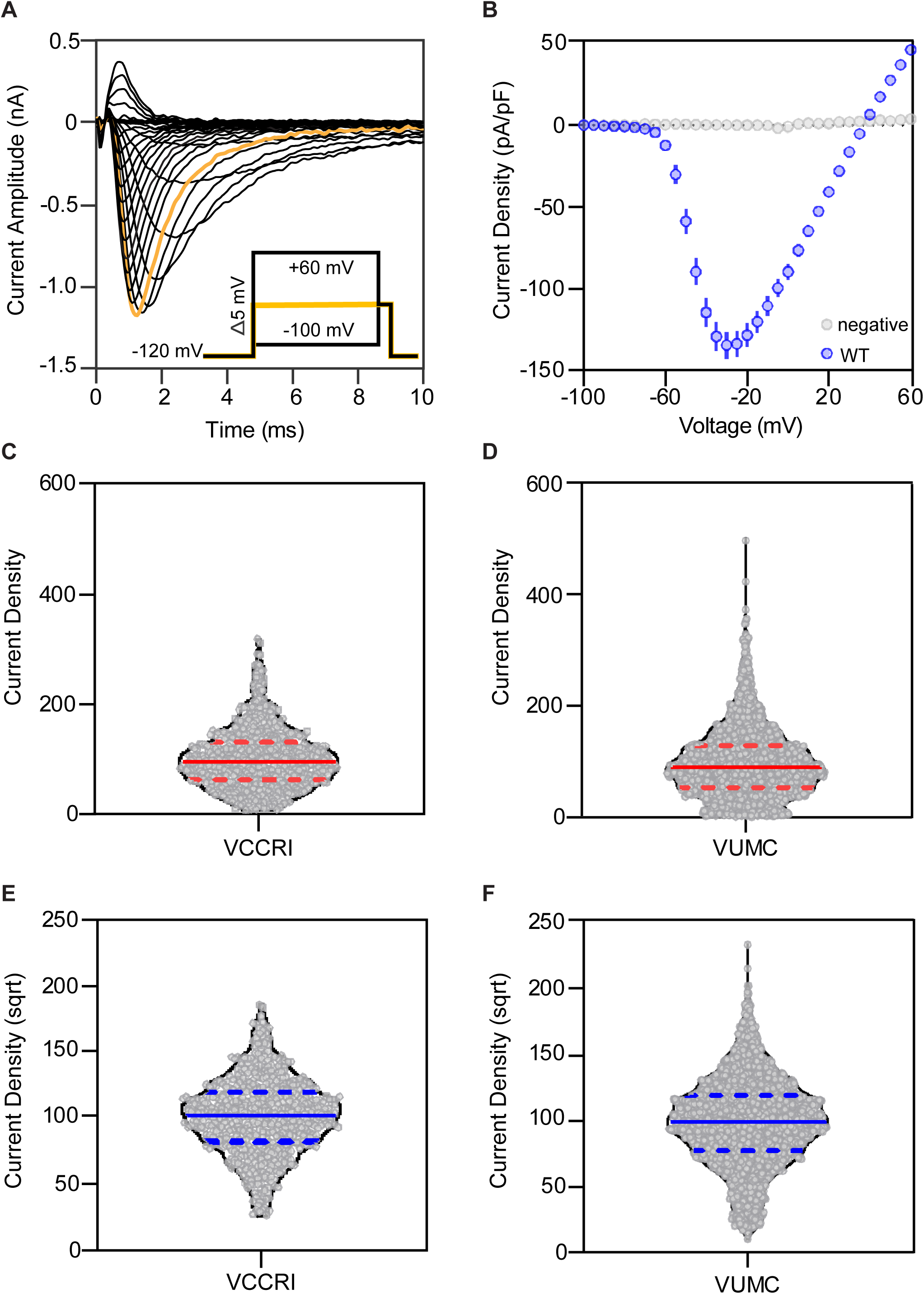
**Establishing an *SCN5A* voltage protocol and data transformation for normal distribution**. (A) Example traces from a single cell expressing WT *SCN5A* illustrating sodium conductance following activation from –100 mV to +60 mV in +5 mV increments. The –30 mV sweep is highlighted in orange and was used for analyses. (B) Current-voltage curves show current densities at voltages –100 mV to +60 mV for WT (blue) and negative cell lines (grey). Mean ± 95% confidence interval values are shown. (C-D) Violin plot of WT *SCN5A* current density at each site (VCCRI N = 609; VUMC N = 2,118 cells). The current density of WT *SCN5A* did not follow a normal distribution (W = 0.956 and W = 0.942 at VCCRI and VUMC, respectively; Shapiro-Wilk test). The median value and 1^st^ and 3^rd^ quartiles are indicated with red lines. (E-F) Violin plots of WT *SCN5A* current density at each site after square root transformation. The square root-transformed current densities more closely followed a normal distribution (W = 0.997 and W = 0.995 at VCCRI and VUMC, respectively; Shapiro-Wilk test). The median value and 1^st^ and 3^rd^ quartiles are indicated with blue lines. The resulting mean ± SD of the square root-transformed distribution was 100 ± 29.2 (VCCRI) and 100 ± 34.1 (VUMC).

### Peak *I*_Na_ density distinguishes benign from pathogenic variants

Many BrS-associated *SCN5A* variants have reductions in peak *I*_Na_ density, the maximum amount of current through the channel.^21^ Z-scores were calculated using the distribution of 25 benign variant controls as reference. Z-scores between –2 and 2 were considered normal, and Z-scores between –2 and –4 were considered moderate LOF, and Z-scores smaller than –4 were considered severe LOF. Normalized current densities for B and P/LP variant controls are shown for VCCRI (Figure 3A) and VUMC (Figure 3B), and the combined VCCRI and VUMC dataset (Figure 3C). Variant-level data are summarized in Tables S5-S7. Across the 49 variants, we observed excellent correlation between sites for peak *I*_Na_ density despite slight differences in cell systems, protocols, and analysis pipelines (Figure 3D; R^2^ = 0.86). Overall, 25/25 B and 23/24 P/LP variant controls had matching results between the two sites for peak *I*_Na_ density, considering this variable as a binary outcome of normal or abnormal. The only variant discordant between the two sites was p.Arg1631His, an LP variant that had reduced *I*_Na_ density in the VCCRI dataset and normal function in the VUMC dataset (Figure 3).

**Figure 3:**
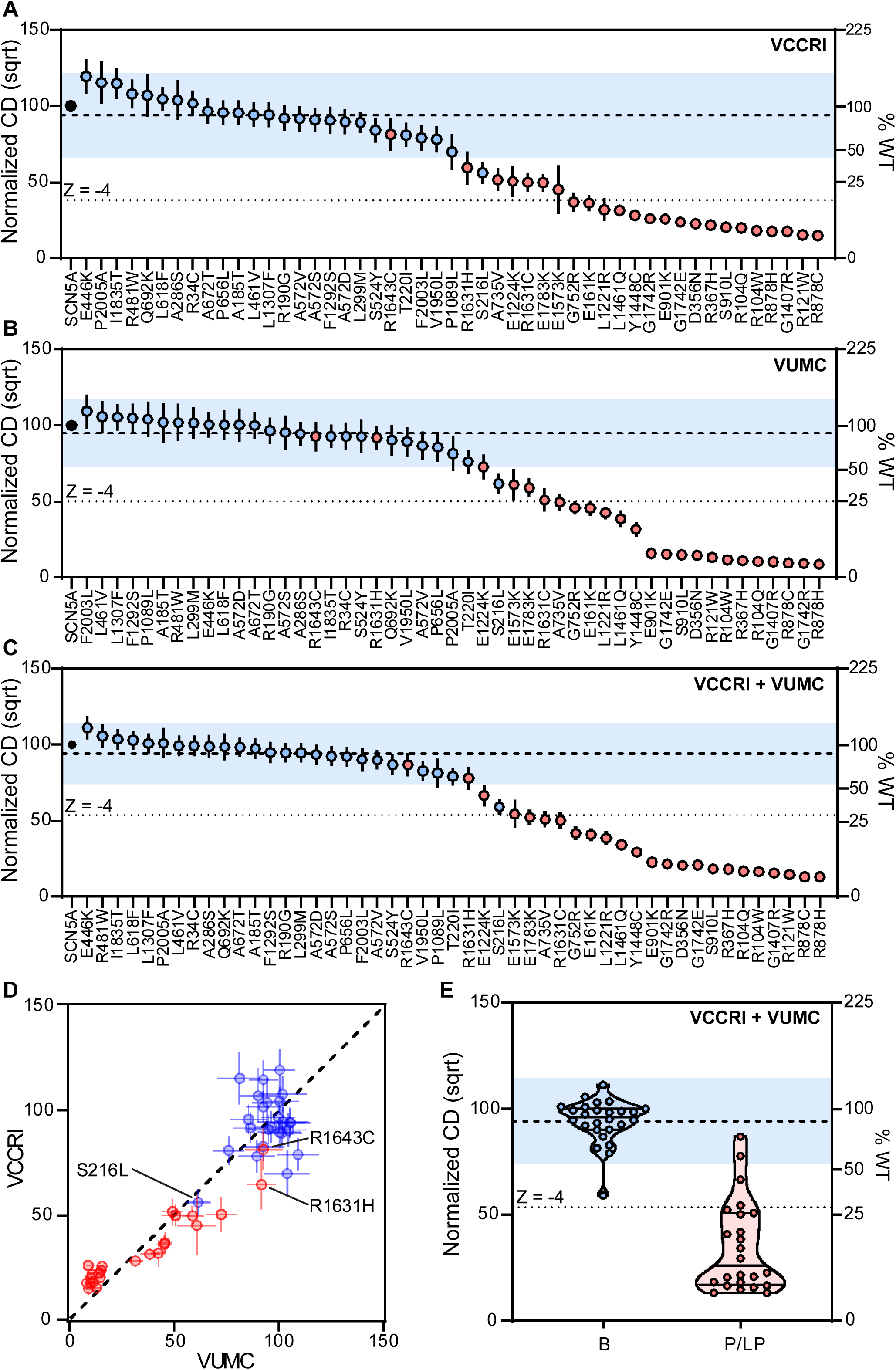
**Distinguishing pathogenic and likely pathogenic variants from benign variants with Na_V_1.5 peak current density measurements**. (A-C) Square-root transformed, normalized peak current densities at –30mV obtained by VCCRI (94.1 ± 13.9; A), VUMC (94.8 ± 11.1; B) and a combined dataset (94.2 ± 10.3; C). *SCN5A* peak *I*_Na_ densities (pA/pF) were measured after holding at –120 mV. The normal functional range was defined as the mean ± 2 SD of the benign variant values (blue region). The peak current measurements distinguished the P/LP and B variants, apart from three variants (p.Ser216Leu, p.Arg1631His, and p.Arg1643Cys) discussed further in the text. (D) Peak current densities from the two sites were highly correlated (p<0.0001; R^2^ = 0.86). (E) Violin plot summary of combined current densities among B and P/LP variants. Blue-filled circles (●) indicated B variant controls. Red-filled circles (●) indicates P/LP variant controls. Data is presented as mean ± 95% CI.

We observed a strong predictive ability of peak I*_Na_*current to distinguish benign from pathogenic variant controls. In the merged dataset (Figure 3C), 22 of 24 P/LP variant controls had moderate or severe LOF (Z < –2). Two P/LP variant controls had current density within normal ranges: p.Arg1643Cys (Z = –0.71) and p.Arg1631His (Z = –1.6), though VCCRI observed partial LOF for p.Arg1631His (Figure 3A). As we describe below, both sites measured abnormal gating properties for p.Arg1631His. In the merged dataset (Figure 3C), 24 of 25 B variant controls were within the normal range of function (Z > –2). One benign variant, p.Ser216Leu, had moderate LOF at both sites and a final Z-score of –3.39. Overall, peak *I*_Na_ density was concordant with ACMG/AMP classifications for 46/49 variants (93.9%).

### Incorporation of additional channel gating parameters

We next studied variant effects on three additional channel gating parameters: steady state activation (SSA), steady-state inactivation (SSI), and recovery from inactivation (RFI) (Figure 4, Supplementary Table 5-7). These parameters quantify the voltages at which channels activate and inactivate and the time needed to recover from inactivation. We observed good to excellent correlation between VCCRI and VUMC for these parameters (SSA R^2^=0.44; SSI R^2^=0.48; RFI T_50_ R^2^=0.91; Figure S2-4). Unlike peak *I*_Na_ density (Figure 3), none of the three gating parameters strongly distinguished B from P/LP controls (Figure 4). We also note that for most variants with LOF – including most P/LP variant controls – these gating parameters could not be measured due to the small *I*_Na_. Therefore, we applied a more conservative Z-score threshold of ±4 for altered effects. Using this cutoff, five P/LP variants (p.Glu1224Lys, p.Ala735Val, p.Glu1783Lys, p.Arg1631His, and p.Arg1631Cys) and no B variants showed altered gating parameters (Table 1, Figure 4). In particular, two variants had extreme changes in RFI ΔT_50_: p.Arg1631Cys (Z = 72.74) and p.Arg1631His (Z = 106.09; Figure 4E-F, H-I, Table 1). This finding resolves the initial discordance between the LP classification for p.Arg1631His and the normal-range peak *I*_Na_ density measured at VUMC. Thus, an integrated analysis of peak current density and other gating parameters yields 23/24 B and 24/25 P/LP variants that have concordant functional assay results and clinical classifications.

**Figure 4:**
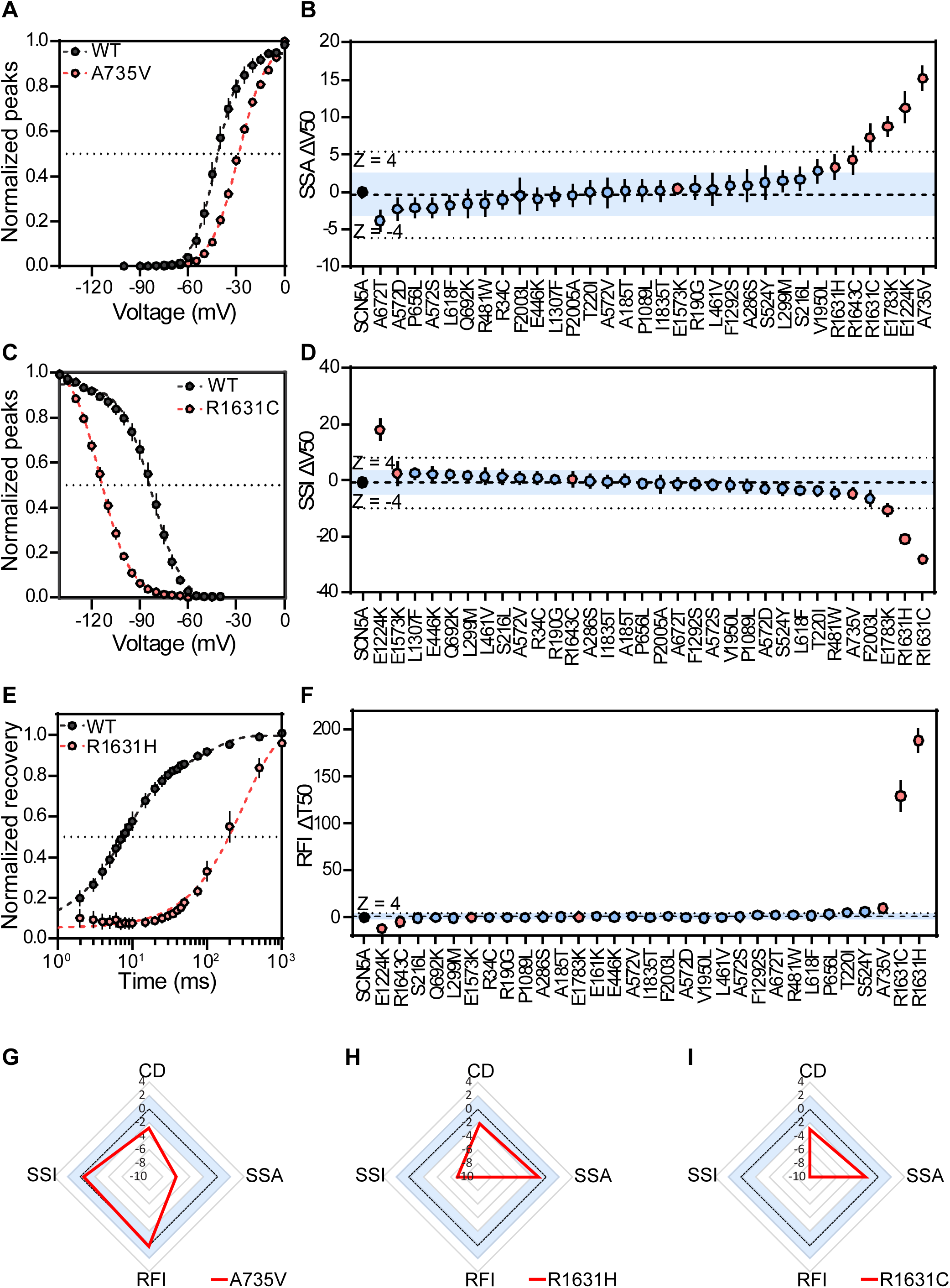
**Analysis of channel gating parameters reveals variants that affect gating**. (A) SSA of p.Ala735Val compared to WT, showing a right-shift in voltage of activation. Boltzmann best-fit curves are plotted. (B) Steady-state voltage of half-activation difference from wildtype (SSA ΔV_50_) for variant controls. Combined SSA ΔV_50_ = –0.37 ± 1.71. ΔV_50_ represents the shift in voltage where variants are half activated, as compared to WT. (C) SSI of p.Arg1631Cys compared to WT, showing a left-shift in voltage of inactivation. Boltzmann best-fit curves are plotted. (D) Steady-state voltage of half-inactivation difference from wildtype (SSI ΔV_50_) for variant controls. Combined SSI ΔV_50_ = –0.37 ± 1.71. ΔV_50_ represents the shift in voltage where variants are half inactivated, as compared to WT. (E) RFI of p.Arg1631His compared to WT, showing a large delay in recovery post inactivation. Double exponential best-fit curves are plotted. (F) RFI difference from WT (ΔT_50_). Combined RFI ΔT_50_ = 0.933 ± 1.77. ΔT50 represents the shift in time required for channels to reach a half-recovered from inactivation state as compared to WT. Many P/LP control variants did not have sufficient peak current to measure the three parameters shown in this figure. (G) Sample radar plots for corresponding variants with extreme shifts in gating parameters: p.Ala735Val (SSA), p.Arg1631Cys (SSI) and p.Arg1631His (RFI). Radar plots for all analyzed variants are presented in Figure S5. Blue-filled circles (●) indicated B variant controls. Red-filled circles (●) indicates P/LP variant controls. In panels B, D, F, G, H and I, the dashed line represents Z = 0, and the blue region indicates Z-scores within ± 2. In panels B, D and F, and the dotted lines indicate Z outside of ± 4. Data is presented as mean ± 95% CI.

**Table 1:** Variants with outlier values for gating parameters.

| Variant | SSA |  | SSI |  | RFI |  | *I*<sub>Na</sub> Density<br>-120 mV hold |  | *I*<sub>Na</sub> Density<br>-90 mV hold |  |
| --- | --- | --- | --- | --- | --- | --- | --- | --- | --- | --- |
|  |  |  |  |  |  |  | Norm |  | Norm |  |
| | $\Delta V_{50}$ | Z | $\Delta V_{50}$ | Z | $\Delta T_{50}$ | Z | (sqrt) | Z | (sqrt) | Z |
| p.Ala735Val | 15.14 | **10.54** | -4.74 | -1.72 | 9.62 | **4.91** | 50.85 | **-4.19** | 43.30 | **-3.18** |
| p.Glu1224Lys | 11.23 | **7.87** | 18.03 | **8.37** | -11.90 | **-7.25** | 66.63 | **-2.66** | 54.98 | **-2.41** |
| p.Arg1631Cys | 7.25 | **5.16** | -28.04 | **-12.04** | 129.60 | **72.74** | 50.28 | **-4.25** | 23.89 | **-4.45** |
| p.Arg1631His | 3.28 | 2.45 | -20.90 | **-8.87** | 188.60 | **106.09** | 77.67 | -1.60 | 39.41 | **-3.43** |
| p.Glu1783Lys | 8.77 | **6.19** | -10.60 | **-4.31** | 0.24 | -0.39 | 52.28 | **-4.05** | 26.62 | **-4.27** |

Severe gating changes ( $Z > +4$ or $Z < -4$ ) were observed in five P/LP variants. p.Arg1631His, which had *I*<sub>Na</sub> density value normal ranges at activation following -120 mV resting potentials, had a hyperpolarized shifts in SSI and severely delayed RFI. This gating effect was also visibly incorporated at activation following -90 mV resting potentials. Bolded numbers indicate measurements outside the benign normal range ( $-4 < Z < 4$ for SSA, SSI, and RFI and $Z < -2$ for *I*<sub>Na</sub> density).

### Peak *I*_Na_ density for depolarization from a –90 mV holding potential

Peak *I*_Na_ density is commonly measured from a holding potential of –120 mV, which ensures that nearly all channels begin the measurement from the closed state. However, under physiological conditions, the resting cardiomyocyte membrane potential is closer to –90 mV.^35^ We hypothesized that variants with abnormal gating properties such as hyperpolarized SSI may have reduced current density when measured from a holding potential of –90mV (Figure 5A). To test this hypothesis, at VCCRI, peak *I*_Na_ densities were measured from a holding potential of –90 mV (Figure 5). Peak *I*_Na_ densities measured from holding potentials of –90 mV and –120 mV were highly correlated (R^2^=0.95, Figure 5B). However, three variants (p.Arg1631Cys, p.Arg1631His and p.Glu1783Lys) had lower *I*_Na_ density when measured from a holding potential of –90 mV compared to –120 mV (Figure 5B). All three of these variants also had hyperpolarized SSI (Figure 4D, Table 1). We next performed square root transformations of the –90 mV peak *I*_Na_ density measurements, and used the mean and standard deviation of benign variant controls to define Z-scores. This single parameter (peak *I*_Na_ density measured from a holding potential of –90 mV) yielded 25/25 B and 23/24 P/LP variant controls that have concordant functional assay results and clinical classifications (Figure 5C). In particular, the measurement at –90 mV peak improved the concordance of the B variant p.Ser216Leu (Z = –3.39 at a holding potential of –120 mV, Z = –1.72 at –90 mV) whilst also increasing the B variant p.Glu446Lys (Z = 1.65 at a holding potential of –120 mV, Z = 2.37 at –90 mV). Evaluation of p.Glu446Lys gating parameters measured in this study does not suggest altered function (Table S5-7). However, as this SCN5A-BrS assay is only validated against LOF, a calibrated GOF assay may be valuable to explore this variant further.

**Figure 5:**
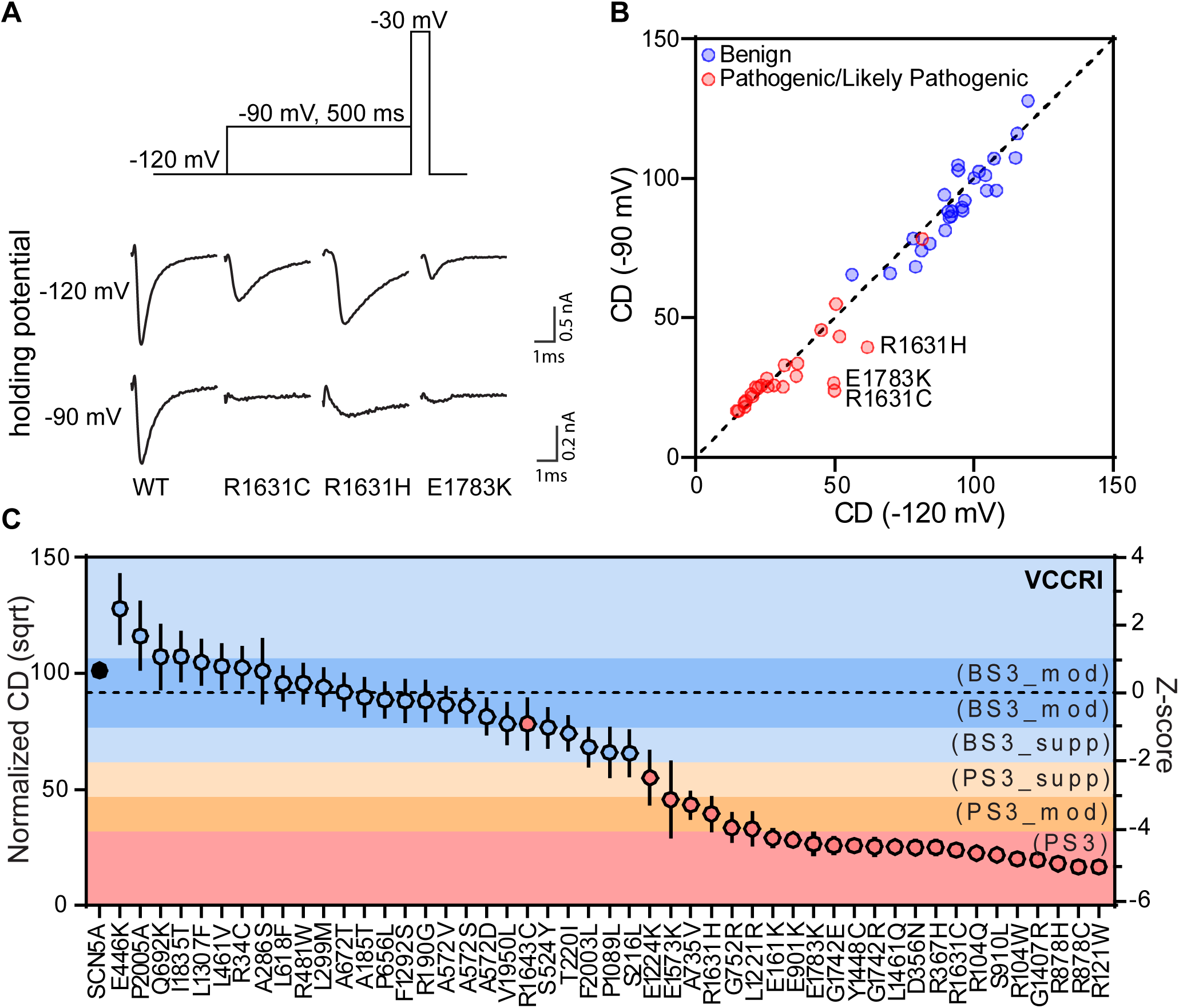
**Current density measured from a physiological resting potential is a single variable that distinguishes benign from pathogenic variants**. (A) Protocol showing holding voltages of –120 mV –90 mV for 500 ms prior to activation of channels. Sample raw traces showing current measured with holding potentials of –120 mV and –90 mV for the same variant cell when depolarized to –30 mV. (B) Correlation between the different holding membrane voltages show strong correlation (R^2^=0.95), with outliers p.Arg1631His, p.Arg1631Cys and p.Glu1783Lys. (C) Normalized current density of variant controls activated from a holding voltage of –90 mV to represent the resting membrane potential of human cardiomyocytes encompasses gating parameters such as SSA, SSI and RFI. Mean = 91.7 ± 14.9. Data is presented as mean ± 95% CI. Blue-filled circles (●) indicated B variant controls. Red-filled circles (●) indicates P/LP variant controls.

### Establishment of evidence strength for functional data

Recommendations by the ClinGen SVI working group^18^ were applied to evaluate the strength of our two *SCN5A*-BrS assay implementations (Figure 6; Table S8). For our first implementation which combined –120 mV holding potential measurements and gating properties, we observed 24/25 concordant B variants and 23/24 concordant P/LP variants (Figure 6A). This yielded OddsPath scores of 24.0 for pathogenic evidence, and 0.043 for benign evidence (Figure 6B). For our second implementation which considered the single parameter of peak *I*_Na_ density from a holding potential of –90 mV, we observed 25/25 concordant B variants and 23/24 concordant P/LP variants. (Figure 6C). This yielded OddsPath scores of 24.0 for pathogenic evidence, and 0.042 for benign evidence. (Figure 6D). In both cases, these values correspond to application of up to PS3_strong (abnormal function) and BS3_strong (normal function) criteria in the ACMG/AMP classification scheme.^8^

**Figure 6:**
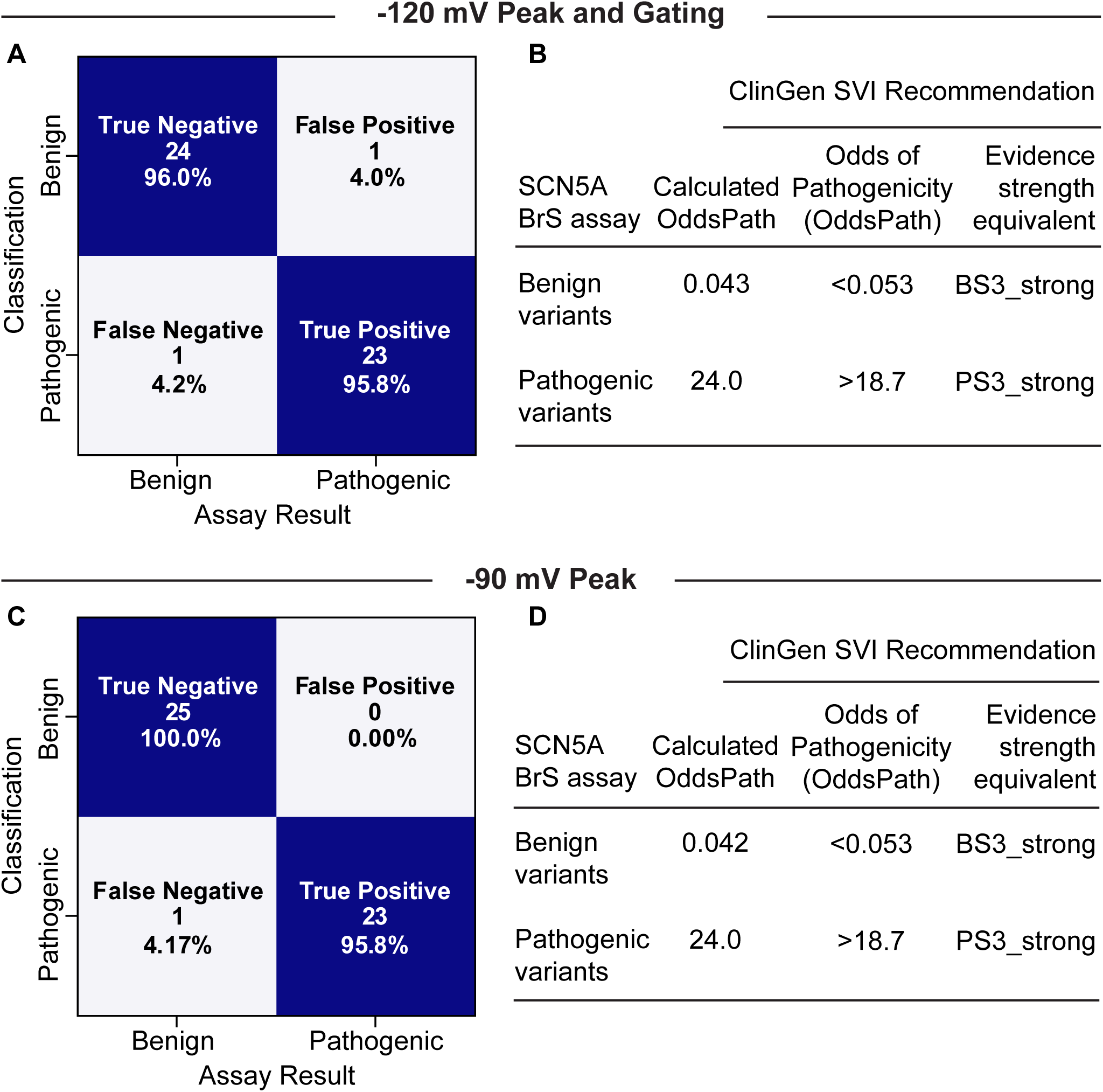
Evaluation of assay according to ClinGen SVI recommendations for functional evidence. (A) Confusion matrix showing sensitivity and specificity of our previously deployed functional assessment combining –120 mV-held peak current (Z = 2) and gating (Z = 4) properties using data from both sites. (B) BS3_strong and PS3_strong may be applied for this combination of parameters functional assay, following ClinGen guidelines ^18^. (C) Confusion matrix showing sensitivity and specificity for a single parameter, –90 mV-held peak current (Z = 2) from the VCCRI site. The B variant p.Ser216Leu is concordantly classified in this implementation of the assay. (D) BS3_strong and PS3_strong may be applied for this single parameter functional assay, following ClinGen guidelines ^18^.

### Functional investigation of clinical *SCN5A* VUS

The calibrated APC assay was used to assess the function of *SCN5A* VUS identified in patients with a clinical diagnosis of BrS or other severe arrhythmia phenotypes associated with *SCN5A* LOF variants.^36^ A list of genes tested for each proband is available in Table S9, additional detected non-*SCN5A* variants are available in Table S10, and classification criteria are available in Table S11. Limited pedigrees with clinical annotations are shown in Figure 7, with the sexes of family members anonymized.

**Figure 7:**
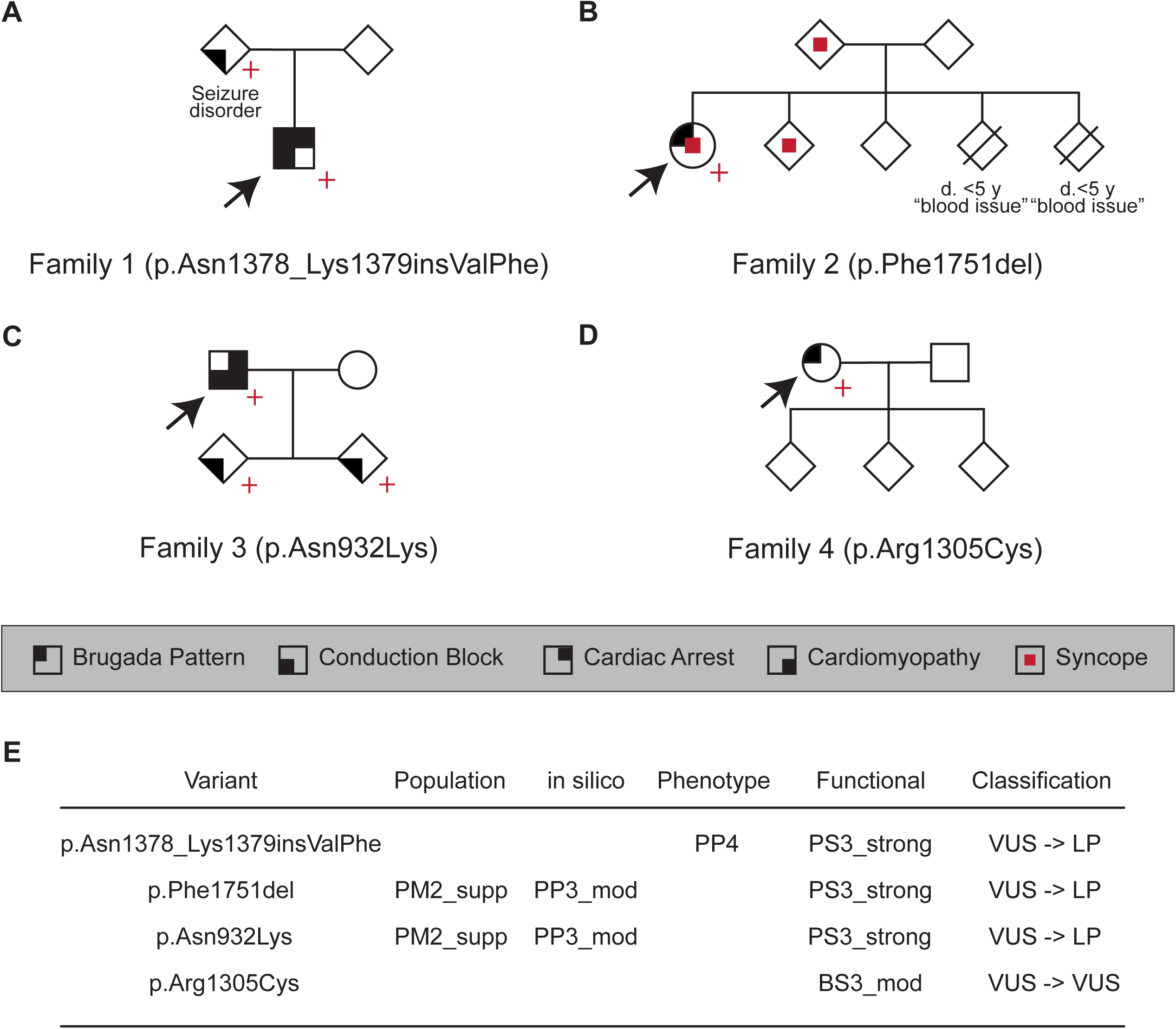
**Reclassification of clinically observed variants with calibrated functional data**. (A-D) Four clinical cases of families segregating *SCN5A* VUS. Arrows indicate the proband. Red crosses indicate variant carriers. The sexes of family members have been anonymized as indicated by diamond-shaped symbols. (A) Proband with sudden cardiac arrest and successful out-of-hospital defibrillation, subsequently diagnosed with Brugada Syndrome and Progressive Cardiac Conduction Defect. The *SCN5A* VUS segregated with conduction disease in the proband and the affected parent. There was a family history of cardiac arrests. (B) Proband with syncope and fever-induced Type 1 Brugada pattern on ECG, with a family history of recurrent syncope, seizures, and one cardiac arrest. Family members were hesitant to engage in genetic testing preventing segregation analysis. (C) Pedigree of family members with conduction block, cardiomyopathy, ventricular tachycardia/fibrillation and aborted cardiac arrest. The *SCN5A* VUS segregated with a severe arrhythmia phenotype in the proband and conduction block in his two young children. (D) Proband with spontaneous Type 1 Brugada pattern on a screening ECG, found to have a VUS in *SCN5A*. (E) ACMG/AMP classification criteria and reclassifications following incorporation of functional data for the four clinically observed variants. Classifications were performed independently at each clinical site by a genetic counselor, clinical geneticist, or physician specializing in genomic medicine.

*Family 1.* The proband was resuscitated from a ventricular fibrillation cardiac arrest during their teenage years. Cardiac workup led to a diagnosis of Brugada Syndrome and Progressive Cardiac Conduction Defect. Genetic testing identified an in-frame insertion in *SCN5A,* p.Asn1378_Lys1379insValPhe (c.4133_4134insTGTGTT), which was classified as VUS. APC revealed that the variant was LOF (Z = –7.62), enabling reclassification to LP after application of PS3_strong (Figure 7). Cascade screening revealed that the *SCN5A* variant was inherited. The proband’s affected parent had a history of a seizure disorder and was subsequently found to have cardiac conduction disease. The parent with the SCN5A variant had a history of a seizure disorder and was subsequently found to have cardiac conduction disease. Additionally, two of the probands’ grandparents (one on each side of the family) had died suddenly in their 30s or 40s.

*Family 2.* The proband was a woman in her 40s who presented after an episode of syncope. Workup revealed fever-induced Type 1 (*i.e.*, definite) Brugada ECG pattern. Genetic testing uncovered the in-frame deletion p.Phe1751del (c.5251_5253del), which was classified as VUS. APC revealed that the variant was LOF (Z = –7.44), enabling reclassification to likely pathogenic after application of PS3_strong. The proband had a family history of recurrent syncope, seizures, and one cardiac arrest. However, besides the proband, the family was hesitant to undergo genetic evaluation, precluding segregation analysis.

*Family 3.* The proband developed complete heart block at a young age and had a 13 second pause, and received an implanted cardioverter defibrillator (ICD). He developed ventricular tachycardia, received shocks from his ICD, and developed heart failure as an adult. Genetic testing identified an *SCN5A* VUS p.Asn932Lys (c.2796T>G). APC revealed that the variant was LOF (Z = –6.52), enabling reclassification to LP after application of PS3_strong. Cascade screening identified the proband’s two young children as variant carriers, and both had a prolonged PR interval (202 msec and 172 msec).

*Family 4.* The proband was a woman in her 60s with an incidental finding of BrS Type 1 ECG and rate-related left bundle branch block during exercise testing. She had no history of syncope and there was no family history of sudden cardiac death. Genetic testing identified an *SCN5A* variant p.Arg1305Cys (c.3913C>T), which has an allele count of 12 in gnomAD v4, and was classified as a VUS. Using APC, we assessed the variant to have normal function (Z = –0.33) and concluded that in this case the *SCN5A* variant was likely not the cause of the BrS ECG pattern. This variant remained classified as VUS by the clinical testing laboratory after incorporation of the functional data (BS3_moderate).

### Reanalysis of previously published APC data

Lastly, we reanalyzed all previously published *SCN5A* APC assay results from our group^17,31,37^ (Table S12). This reanalysis used the same analysis protocol described in this paper, including features like square root transformation of current densities and Z-score-derived cutoffs that were not implemented in our previous studies. We also assigned recommended functional evidence BS3/PS3 strengths to these variants using our updated analysis framework (Table S12).

## Discussion

This study describes an *SCN5A* functional assay that was independently tested and calibrated by two research centers to facilitate *SCN5A-*BrS variant classification. Significant benefits of this assay include replicability across sites, the ability to distinguish B and P/LP variant controls following ClinGen guidelines, and implementation of Z-scores to quantitatively interpret VUS. We also demonstrate a single parameter (peak current measured from a physiological holding potential of –90 mV) that can accurately distinguish B from P/LP controls.

### Multi-site validation of a *SCN5A*-BrS patch-clamp assay

Independent data generation and analyses were performed at two independent sites with similar experimental and analysis pipelines. To determine the reproducibility of the *SCN5A* APC assay, each site independently tested all variant controls. Between the two sites, we observed strong correlations for peak current density (R^2^ = 0.86) and good to excellent correlation for the other gating parameters. For peak *I*_Na_ density, 48/49 variants had concordant results between the two sites (considering each variant as having normal or abnormal function). Future functional assays may benefit from testing at independent sites to demonstrate reproducibility prior to clinical use.

### The calibrated assay can discriminate between variant controls

We describe two implementations of the assay, one that integrates peak *I*_Na_ density and gating measurements, and a second that uses a single parameter (peak *I*_Na_ density measured from a physiological holding potential). Both implementations yielded concordant results in nearly all variant controls (24/25 B and 23/24 P/LP for implementation 1, 25/25 B and 23/24 for implementation 2). After OddsPath calculations as recommended by the ClinGen SVI working group,^18^ both implementations of the assay attained strong levels of evidence for both PS3 (abnormal function) and BS3 (normal function).^8^

Two variant controls had discordant results with their clinical classifications in the peak *I*_Na_ and gating assay, p.Ser216Leu and p.Arg1643Cys. The B variant p.Ser216Leu had partial LOF in peak *I*_Na_ (Z = –3.39 with holding at –120 mV). In the –90 mV holding potential measurement, the Z-score was increased to Z = –1.72 and fell within the normal-function range (Z within ± 2). A previous report also measured partial LOF for this variant in HEK293 cells.^38^ In addition, this variant has also been reported to have late current in HEK293 cells.^39^ p.Ser216Leu is present in the population at a much higher than expected frequency for a highly penetrant disease-associated variant (allele frequency > 0.05% in gnomAD).^40^ However, p.Ser216Leu has also been observed in multiple patients with arrhythmia phenotypes, including BrS,^38,41,42^ Type 3 Long QT syndrome,^43,44^ Sudden Infant Death Syndrome,^39,45,46^ and atrial fibrillation.^47,48^ Given the multiple arrhythmia reports and the borderline partial LOF phenotype of the variant, p.Ser216Leu may be a risk allele for arrhythmia phenotypes, analogous to *KCNE1* p.Asp85Asn.^49^ The LP variant p.Arg1643Cys also had a discordant outcome in our assay, with a functionally normal range peak current (Z = –0.71 and Z = –0.88 with holding potentials of –120 mV and –90 mV, respectively). The only aberrant functional phenotype was a depolarized shift in SSA (+4.31 mV, Z = 3.15), consistent with previous findings.^50^ p.Arg1643Cys has been previously reported in multiple BrS cases^50–53^ and we classified it as LP (PS4_moderate, PM5, PM2_supporting, PP3). Although p.Arg1643Cys was absent from gnomAD v2.1.1 and v3.1.2 used in this study, it is present in gnomAD v4 (allele count of 4/86,258 alleles in the South Asian population). Future work could further investigate the functional impact of p.Ser216Leu and p.Arg1643Cys on channel function in other contexts and cell models (e.g. cardiomyocytes) and help further clarify their disease risk.

### Incorporation of gating changes as well as current density measurements

Peak current is the strongest molecular correlate of BrS and has been associated with risk of events and disease penetrance.^21,54^ Most patch clamp studies of *SCN5A* have used a resting membrane potential of –120 mV in activation protocols to ensure that nearly all channels were closed at the start of the measurement. However, this configuration might overestimate the available peak *I*_Na_ for BrS-associated *SCN5A* variants that affect Na_V_1.5 gating. Some studies have also evaluated the impact of using a resting potential of –90 mV, which is similar to the resting potential of human cardiomyocytes.^55–57^ The present study examined peak current density for variants at resting potentials of both –120 mV and at –90 mV.

When compared to a holding membrane potential of –120 mV, three variants (p.Arg1631Cys, p.Arg1631His and p.Glu1783Lys) had relatively greater reduction in current density when measured with a holding membrane potential of –90 mV (Table 1; Figure 5). These three variants also had altered gating properties in our study (Table 1) and previous reports.^17,55–61^ Altered gating can impact current density by reducing channel availability. Hyperpolarizing shifts in SSI can lead to channels that are not completely inactivated at physiological membrane potentials (and therefore also cannot RFI) and delayed RFI can weaken subsequent activations. As such, holding the resting membrane potential at –90 mV offers more physiologically-relevant functional evaluations while holding membrane potentials at – 120 mV enable the scrutiny of gating parameters that may underlie the abnormal function detected at –90 mV.

### Translational Impact of *SCN5A* APC Assays

Our validated assay was used to assess the function of *SCN5A* variants observed in patients with suspected Brugada Syndrome or other severe arrhythmia phenotypes through collaboration with their primary medical teams. After functional data were provided to each clinical care site, local multi-disciplinary teams performed variant reclassification according to their interpretations of the ACMG framework. Three of the four examined VUS had abnormal function, and addition of this functional data resulted in all three of these variants being reclassified to LP. We anticipate that our calibrated assay will be used in the future to characterize additional *SCN5A* VUS detected in cases of suspected BrS or other arrhythmias associated with *SCN5A* LOF.

### Limitations

*SCN5A* is associated with various other cardiac phenotypes besides BrS, such as long QT syndrome, dilated cardiomyopathy, familial atrial fibrillation, and sick sinus syndrome.^2,58,62,63^ Although our APC assay could likely be used to detect other types of Na_V_1.5 dysfunction, this study only examined the *SCN5A*-BrS relationship linked to Na_V_1.5 LOF variants.^7^ To enable high-throughput studies, HEK293 cell lines were employed for experiments rather than induced pluripotent stem cell-derived cardiomyocytes or animal models. These models may be more physiologically accurate but are more costly and at the present are lower throughput than heterologous expression experiments.^64^ *SCN5A* function can be influenced by several features not modelled in our study, including alternative spliced isoforms,^65^ or expression of additional genes such as β-subunits^39,65,66^ or transcription factors.^5^ Despite these limitations, the assay was still able to accurately distinguish benign from pathogenic variant controls.

## Future Directions

Additional known and yet to be discovered *SCN5A* VUS could be studied using our calibrated functional assay. Further investigations of the two variants with discordant assay results may reveal new biological mechanisms. Further implementation of multi-site replication and calibration for additional genes and disorders will broaden the impact of APC assays for precision medicine.

## Declarations

## Data Availability

All data produced in the present study are available upon reasonable request to the authors.

https://github.com/GlazerLab/

https://github.com/VCCRI/

## Acknowledgements

We thank Kenneth Matreyek and Doug Fowler for the HEK293 landing pad cells, and Lorena Harvey, Maria Calandranis, and Paige Roberson with *SCN5A* cloning assistance and cell preparation. Figure 1 was created with BioRender.

## Authors’ contributions

JM, MO, JV, CN and AG contributed to the original research and analyses, drafting, and editing this paper. EB, KT, JI, AM, JS, GD, KA, BS, AS, BF, KD, VP, HC, MP and DR contributed additional genetic and clinical expertise, and helped edit the paper. All authors approved the final manuscript.

## Ethics approval and consent to participate

The University of Washington Human Subjects Division, Institutional Review Board, BioMedical Committee B reviewed and granted approval for the BBI-CVD study # 14771. The University of Washington’s IRB FWA # is #00006878. The IRB granted expedited approval for the study with consent, HIPAA authorization, parental permission and assent waived, and involvement of children was approved. Stanford University School of Medicine IRB gave ethical approval, and patient consent was waived based on removal of all PHI. Vanderbilt University Medical Center IRB granted approval #9047. Additionally, signed informed consent was received from patient. Clinical case from The Royal Melbourne Hospital does not have IRB/REC as it is not an enrolled patient however, patient consent for publication was received.

## Availability of data and materials

All functional data are available in the accompanying Supplement and Supplementary files in .csv format. Code used to process these files is available at https://github.com/GlazerLab/ and https://github.com/VCCRI/. Individual clinical data beyond those presented in the manuscript are withheld for patient privacy. Variant classifications and functional data will be uploaded to ClinVar upon publication.

