## Supplemental methods and figures for "Multi-site validation of a functional assay to adjudicate *SCN5A* Brugada Syndrome-associated variants"

#### **Site Specific Protocols**

##### VUMC

*Cloning of Variant Constructs.* *SCN5A* plasmids were manually cloned using a previously described ‘Zone’ system (1). In brief, *SCN5A* cDNA was split among small expression plasmids that underwent mutagenesis with the QuikChange Lightning Mutagenesis kit as previously reported (Agilent). All primers used are in Table S4. Variant zone plasmids underwent subcloning by restriction digest into a AttB:*SCN5A*:IRES:mCherry-blasticidinR plasmid.

*Transfection of Cells.* All Vanderbilt EP data was obtained from HEK293T “negative selection” landing pad (LP) cells (kind gift of Kenneth Matreyek). By combining these cells with the plasmids above, we obtained a single integration per cell and a consistent level of expression as determined by mCherry. *SCN5A* plasmids were stably integrated using transposase, followed by negative and positive selection.

*High-throughput Electrophysiology.* All variant cell lines were studied in at least 2 independent transfections in unique SyncroPatch experiments. All electrophysiology was conducted on the SyncroPatch 384 PE with identical solutions and cell preparation as previous reports (1). All protocols to obtain peak current, voltage of half activation, voltage of half inactivation, and time of 50% recovery from inactivation are presented in Figure S1. Internal solution (mM): 10 Na, 20 Cl, 110 F, 120 Cs, 10 HEPES, 10 EGTA. External solution (mM): 80 Na, 106 Cl, 1 Mg, 4 K, 2 Ca, 10 HEPES, 5 Glucose.

*Data-analysis*. SyncroPatch data were analyzed as previously reported. We obtained peak current at -120 mV holding voltage by using our activation protocol at -20 mV and dividing by cell capacitance. Peak current from a holding voltage of -90 mV was similarly obtained from our inactivation protocol, by dividing peak current at -20 mV by cell capacitance. All peak current densities were normalized to WT current densities across each cell and then square root transformed obtain normally distributed data.

##### VCCRI

Flp-In *SCN5A* HEK293 cell lines were generated using methods we previously published for *KCNH2* (2).

*Cloning of SCN5A Constructs.* WT *SCN5A* cDNA in pcDNA^TM^5/FRT/TO (Thermo Fisher Scientific cat # V652020) was designed to have eight segments separated by restriction sites (SbfI, PmII, NheI, BamHI, KpnI, BstEII, NsiI) for subcloning purposes. Two restriction sites were introduced (PmII, NsiI), one restriction site was removed (BamHI), and Q1077 was removed to produce the WT *SCN5A* ΔQ1077 cDNA where it was used as the template for generating variant plasmids by Genscript Inc. (Piscataway, USA).

*Transfection of Cells.* *SCN5A* variant plasmids were transfected into HEK293 cell lines at VCCRI using pOG44 Flp-Recombinase Expression Vector (Thermo Fisher Scientific, cat. # V600520) and Lipofectamine 3000 (Thermo Fisher Scientific, cat. # L3000015), per product protocol. These stable cell lines were selected for hygromycin-resistance and maintained using DMEM (Thermo Fisher Scientific, cat. # 10566016) supplemented with 10% FBS (Sigma-Aldrich, cat. # 12007C), 10 ug/mL Blasticidin (InvivoGen, cat. # ant-bl-1) and 200 ug/mL Hygromycin (Thermo Fisher Scientific cat. # 10687010).

*High-throughput Electrophysiology.* All variant cell lines were harvested, as previously published (2), using Accumax (Sigma-Aldrich, cat. # A7089). Patch clamp experiments include 10 *SCN5A* variants, WT *SCN5A* for positive control, and a non-transfected HEK293 cell line as negative control. All variants were studied in technical duplicates. Experiments were conducted on the SyncroPatch 384 PE to measure conductance using specialised voltage protocols. Internal solution (mM): 10 Na, 10 CsCl, 110 CsF, 10 HEPES, 10 EGTA; pH 7.2 with CsOH. External solution (mM): 20 Na, 5 KCl, 1 Mg, 2 Ca, 120 TEACl, 10 HEPES, 5 Glucose; pH 7.4 with NaOH.

*Data-analysis*. SyncroPatch data were analysed using a MATLAB script specifically designed for *SCN5A* data analysis. Peak current densities, normalised to capacitance (pA/pF), were obtained at -20mV following holding potentials of -120mV for complete current densities and -90mV for physiological current densities. Data is square root transformed for Gaussian distribution then normalised to mean WT per experimental plate to ensure uniformity of data between experiments. The outcomes of the analysis include calculation of peak current density under non-physiological and physiological voltages, voltage at half activation and inactivation during steady-state (V_50_), and time at half recovery from inactivation (T_50_).

#### **Variant selection**

We originally selected a total of 51 control variants: 27 benign variants, 19 likely pathogenic variants, and 5 pathogenic variants. However, two benign variants (p.Ser1903Leu and p.Ser1786Asn) technically failed - one during the cloning stage at VUMC and one during the transfection stage at VCCRI. We excluded these variants from the calibration analyses since we could not measure them at both sites.

#### **Site specific QC parameters**

|  | **VUMC** | **VCCRI** |
| --- | --- | --- |
| R_seal_ | > 500 MΩ | > 500 MΩ |
| C_slow_ | 5-25 pF | 3-30 pA/pF |
| R_Series_ |  | < 50 MΩ |
| Leak corrections |  | ±40 pA from baseline |
| Time to peak |  | 1.1 ms at 0mV |
| Outlier removals | > 3 SD |  |
| Current for SS analysis | > 100 pA | 400 to 2000 pA |
| Current for RFI analysis | > 100 pA | > 1000 pA |
| Chip used | Medium resistance | Low resistance, large hole |

#### **Equations used in APC Analysis**

Calculations were implemented in R (VUMC) and MATLAB (VCCRI) during analysis.

Current Density:

$$Current density (pA/pF) = conductance / capacitance$$

Boltzmann equation:

$$Boltzmann equation = I_{Max} + ( I_{Max} - I_{Min} ) / (1+{exp}^{(V50-x)/k} )$$

where, $I_{Max}$ is the maximum peak current, $I_{Min}$ is the minimum peak current, $V50$ is the voltage required for half maximal activation, $k$ is the slope factor.

Steady State Activation:

$$INa = GNa * (V-Vrev)$$

Where $INa$ is the peak sodium current during depolarisation, $GNa$is the conductance, $V$ is the voltage at which depolarisation occurs, $Vrev$ is the reversal potential, and ${INa}_{Max}$ is the maximum peak sodium current during depolarisation.

Steady-state Inactivation:

$$INa = GNa - {GNa}_{Max}$$

Recovery from inactivation:

$$RFI = Y0+\left( \left( Plateau-Y0 \right)*\left( 100-PercentFast \right)*.01 \right)*\left( 1-\exp\left( -KSlow*x \right) \right)$$

$$+ ((Plateau-Y0)*PercentFast*.01)*(1-exp(-KFast*x))$$

Z-score calculations

$$Z = \frac{Raw value - Mean(Benign variant values)}{Standard Deviation \left( Benign variant values \right)}$$

Sample size calculation:

To detect 25% difference at 90% power, where α = 0.05

$$k= \frac{n^{2}}{n^{1}}=1$$

$$n_{1}= \frac{{\left( \sigma_{1}^{2}21+\frac{\sigma_{2}^{2}}{K} \right)\left( z_{1}-\frac{\alpha}{2}+z_{1}-\beta\right)}^{2}}{\Delta^{2}}$$

$$n_{1}= \frac{{\left( {30}^{2}+\frac{{30}^{2}}{1} \right)\left( 1.96+1.28 \right)}^{2}}{{25}^{2}}$$

$$n_{2}=K*n_{1}=30$$

Δ = |μ_2_-μ_1_| = absolute difference between two means
σ_1_, σ_2_ = variance of mean #1 and #2
n_1_ = sample size for group #1
n_2_ = sample size for group #2
α = probability of type I error (usually 0.05)
β = probability of type II error (usually 0.2)
z = critical Z value for a given α or β
k = ratio of sample size for group #2 to group #1

### **Supplementary Figures**

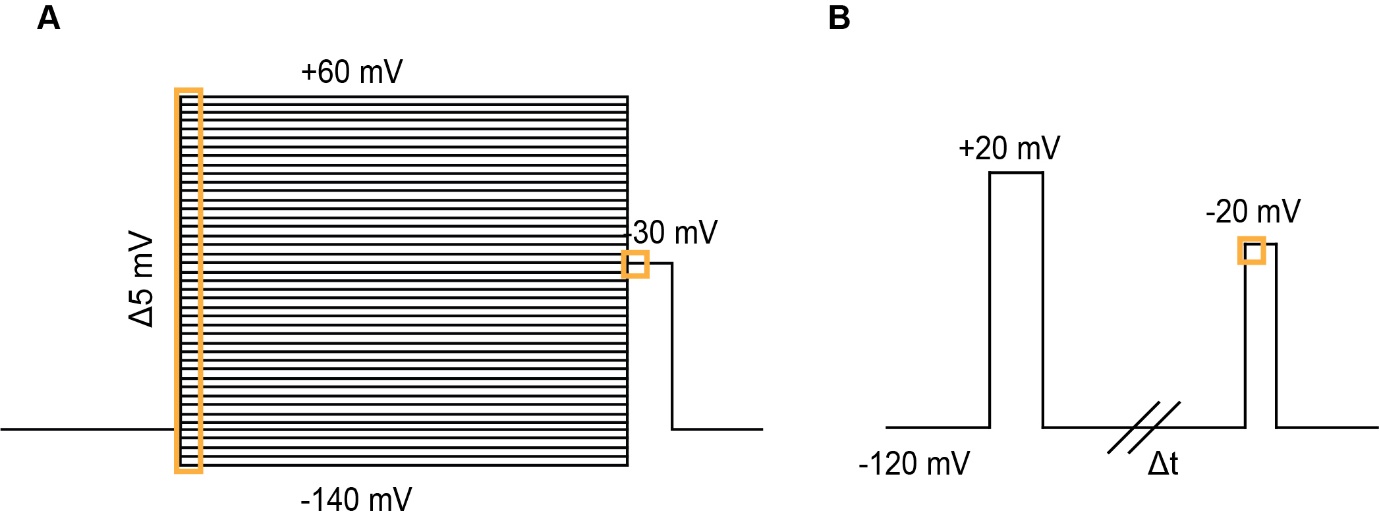

#### **Figure S1: Voltage protocols**

**(A)** Peak *I*_Na_ density measurements and steady-state activation was obtained at various voltages up to +60 mV, Δ5 mV, and steady-state inactivation was measured at -30 mV following each activation. **(B)** Recovery from inactivation is obtained from a -20 mV test-pulse which follows a pre-pulse at +20 mV with increasing time intervals between the two pulses (Δt). All recovery data is normalized to a control pulse of -20 mV at the start of the voltage protocol.

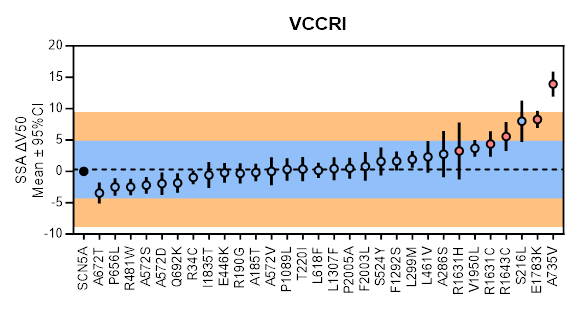

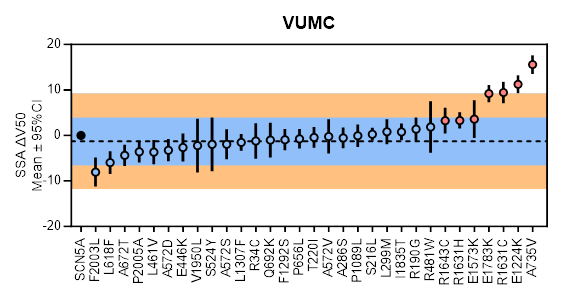

#### **Figure S2: Steady-state activation (-120 mV holding potential)**

Changes in steady-state activation (SSA ΔV_50_) measured at each site. (A) VCCRI (0.289 ± 2.29). (B) VUMC (-1.19 ± 2.24). Mean indicated by dashed line. Blue region indicates ± 2SD. Orange region indicates + 4SD. Blue-filled circles (●) indicated B variant controls. Red-filled circles (●) indicates P/LP variant controls.

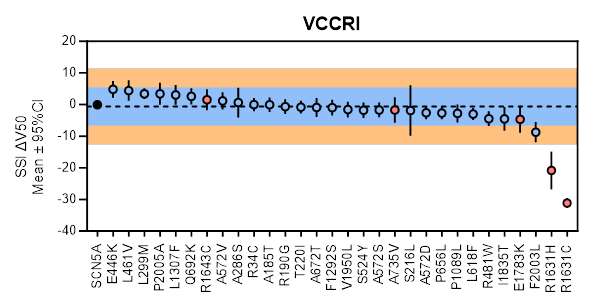

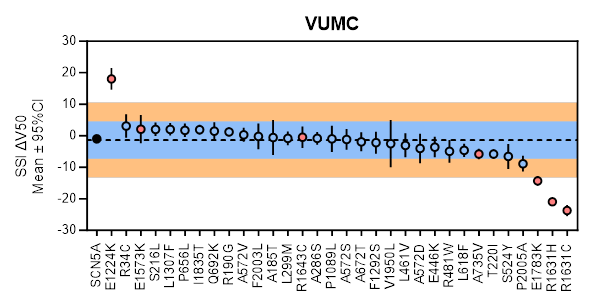

#### **Figure S3: Steady-state inactivation (-120 mV)**

Changes in steady-state inactivation (SSA ΔV_50_) measured at each site. (A) VCCRI (-0.60 ± 3.02). (B) VUMC (-1.35 ± 2.96). Mean indicated by dashed line. Blue region indicates ± 2SD. Orange region indicates + 4SD. Blue-filled circles (●) indicated B variant controls. Red-filled circles (●) indicates P/LP variant controls.

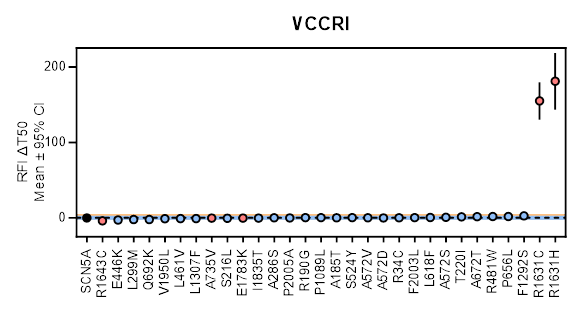

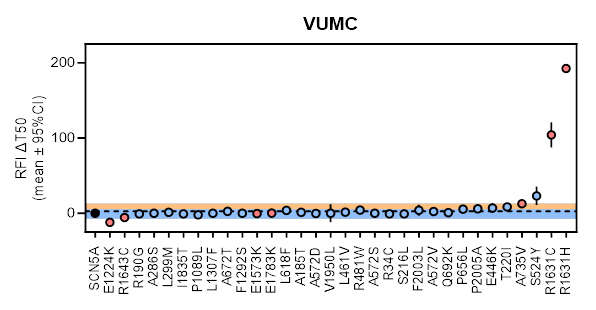

#### **Figure S4: Recovery from inactivation (-120 mV holding potential)**

Changes in recovery from inactivation (RFI ΔT_50_) measured at both sites uncovered significantly prolonged recovery in variants R1631C and R1631H. (A) VCCRI (0.217 ± 1.26). (B) VUMC (2.64 ± 5.01). Mean indicated by dashed line. Blue region indicates ± 2SD. Orange region indicates + 4SD. Blue-filled circles (●) indicated B variant controls. Red-filled circles (●) indicates P/LP variant controls.

#### **Figure S5: Radar plots to integrate gating parameters (-120 mV)**

Gating changes for all variants were measured following a hold of -120 mV. Z-score scale used to for comparison. Blue shade (between Z=2 and Z=-2) indicates functionally normal region as determined by the benign variants. Benign variants in blue. P/LP variants in red and where variants did not produce sufficient current for analysis, only markers for available parameters shown.

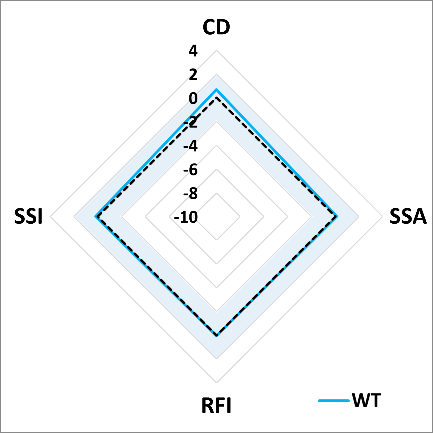

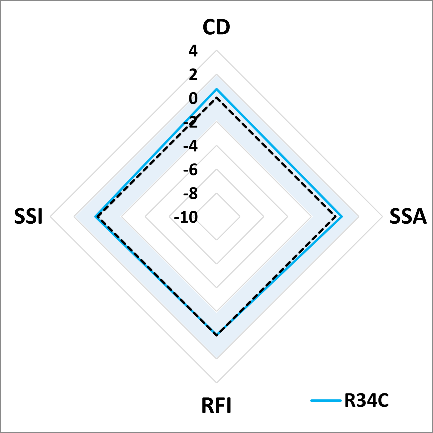

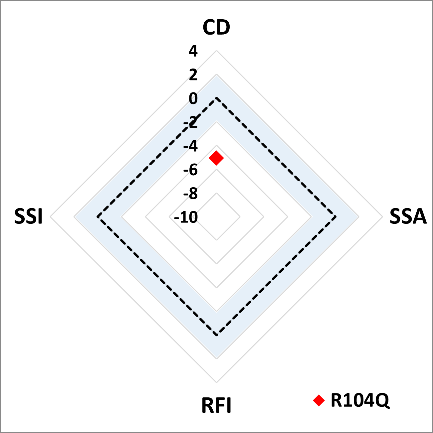

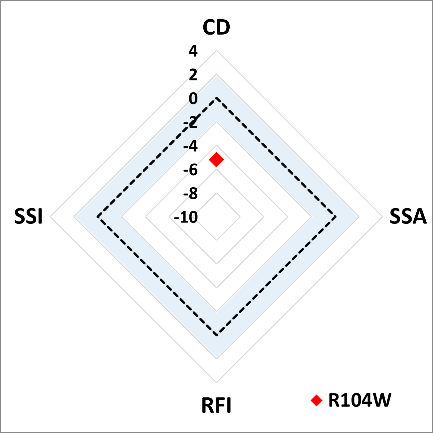

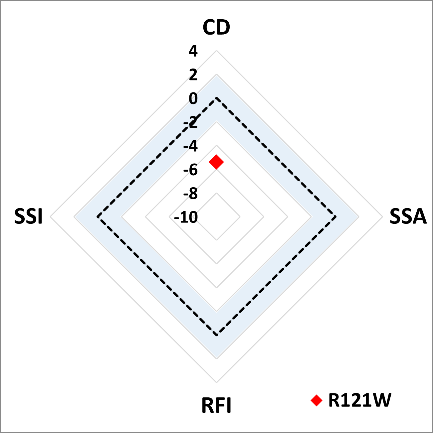

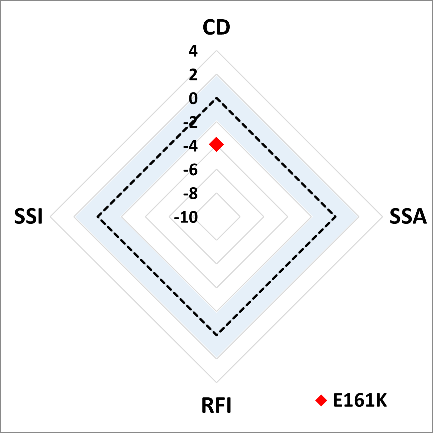

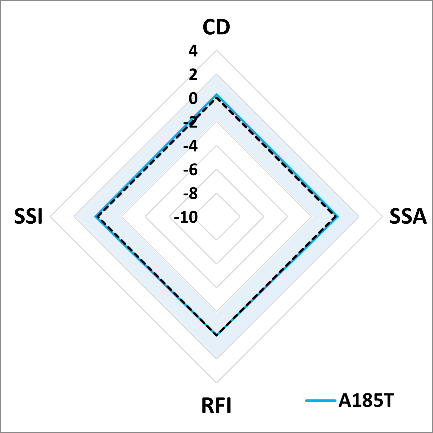

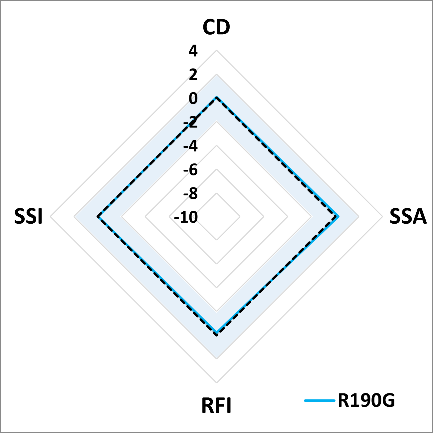

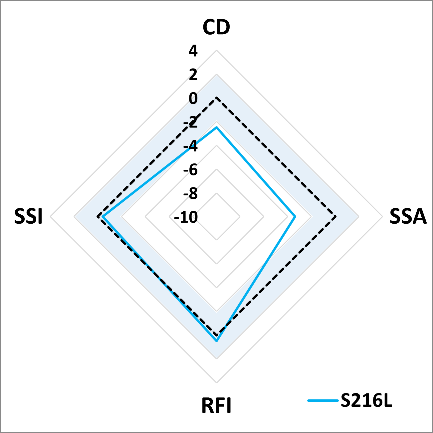

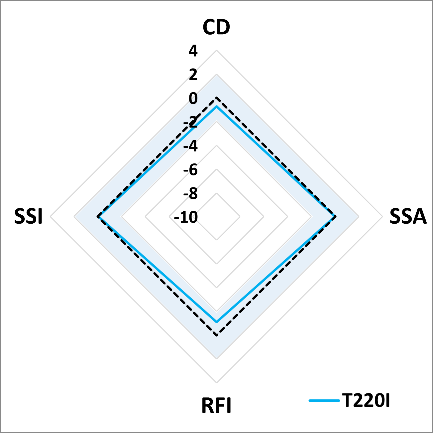

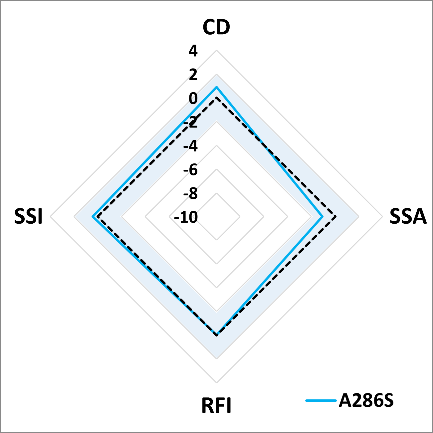

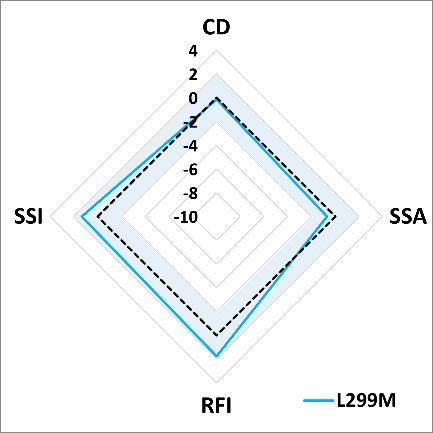

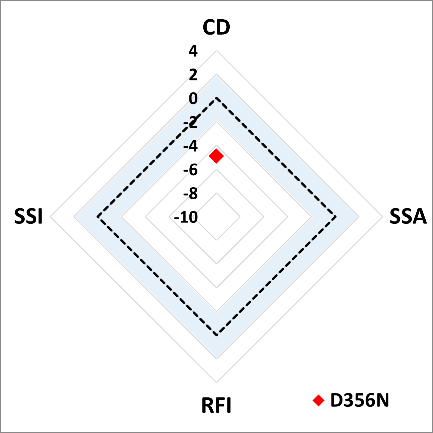

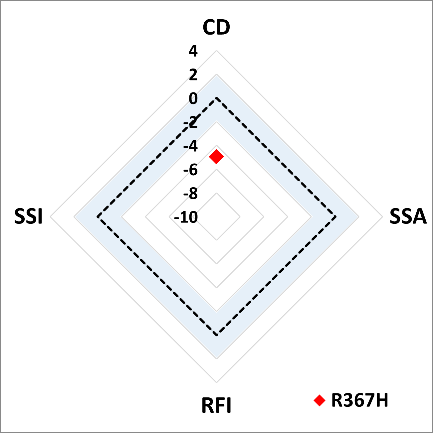

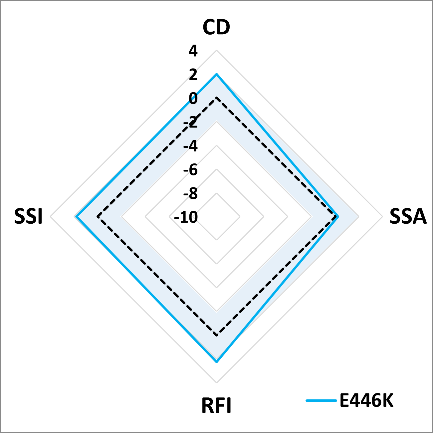

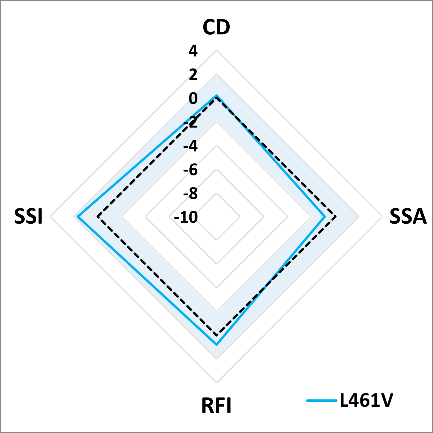

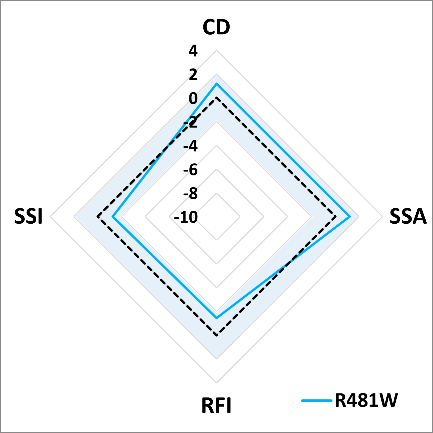

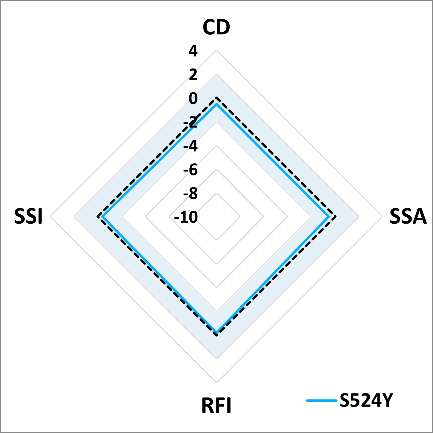

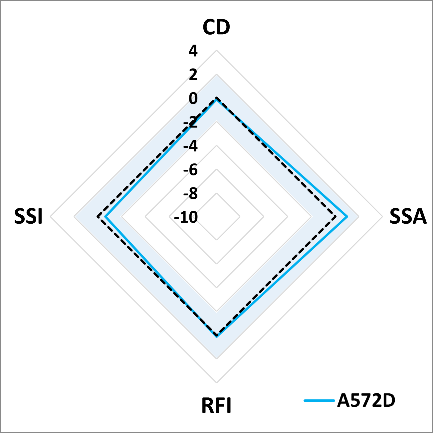

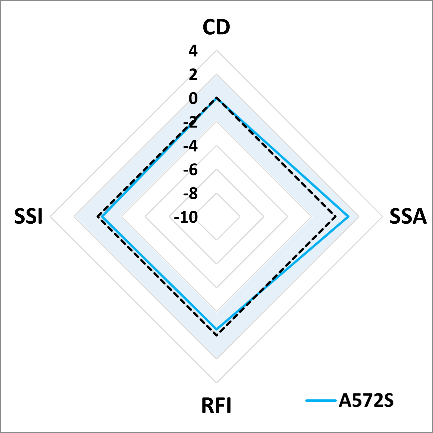

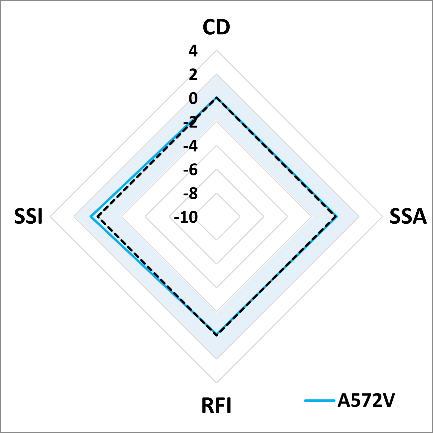

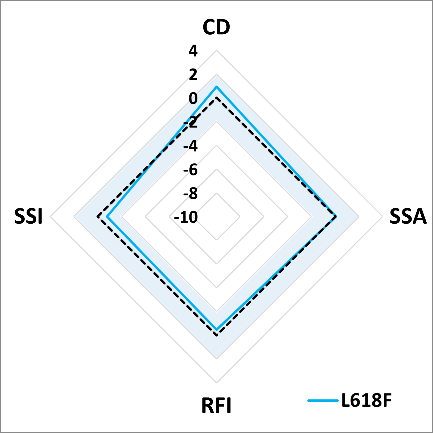

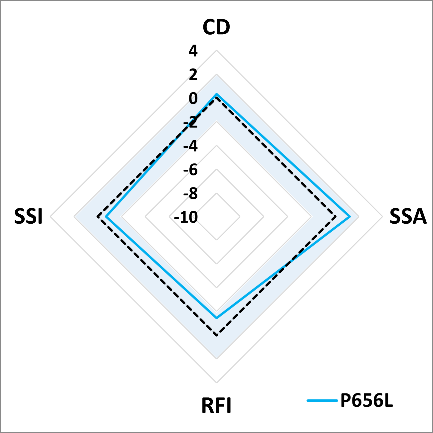

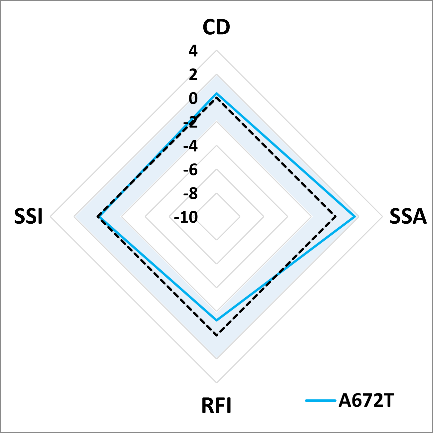

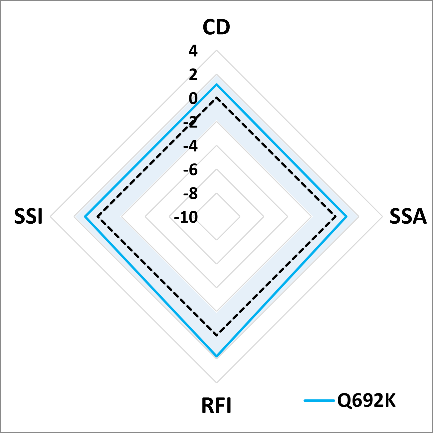

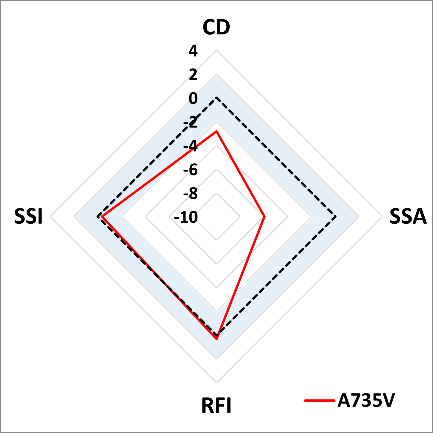

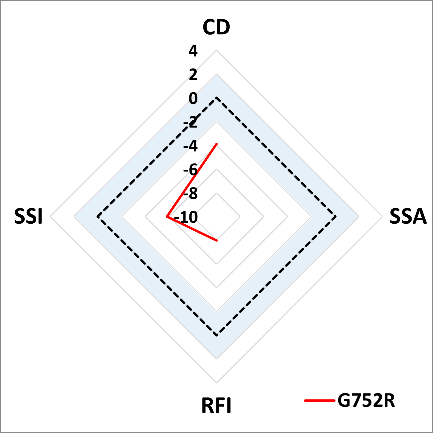

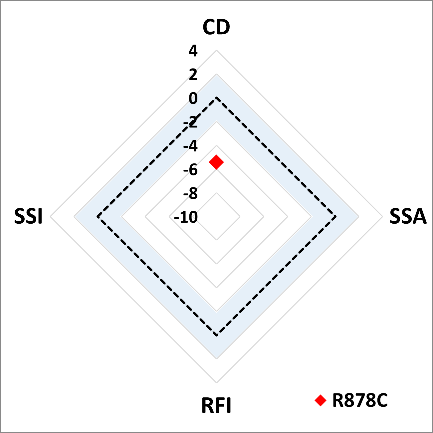

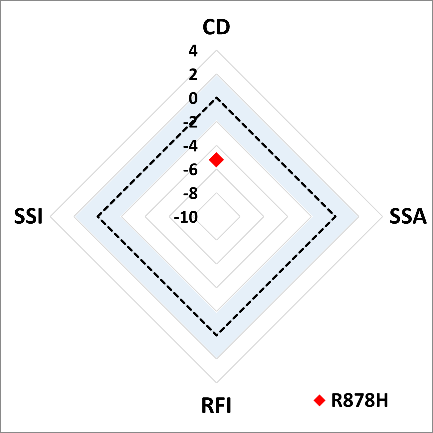

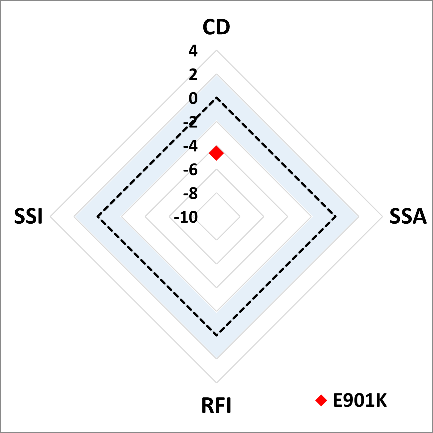

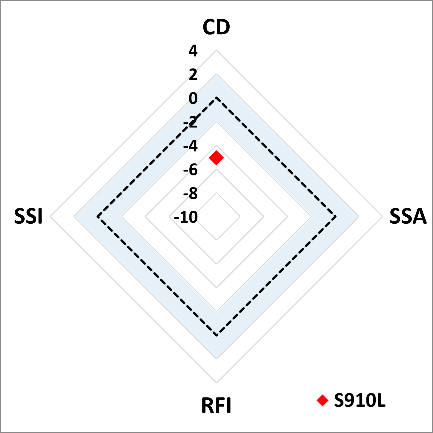

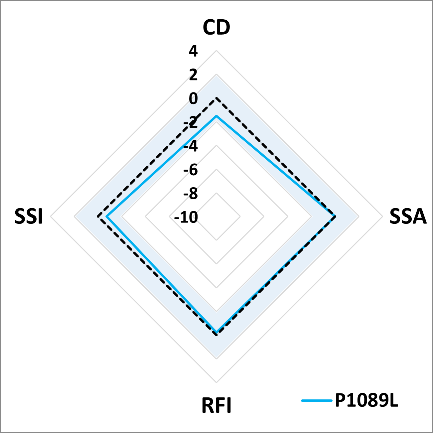

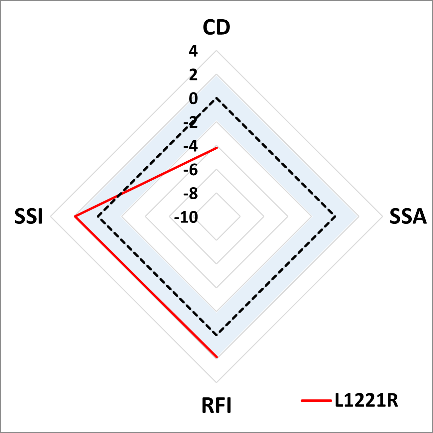

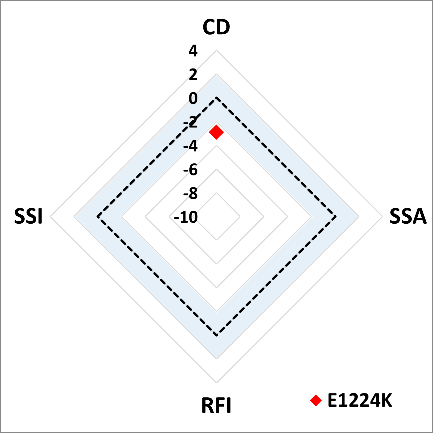

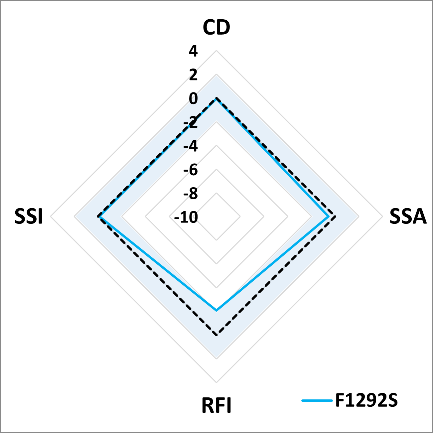

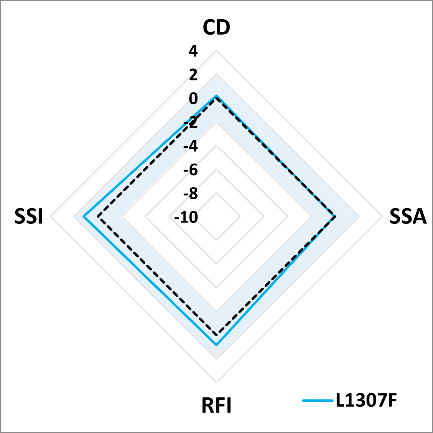

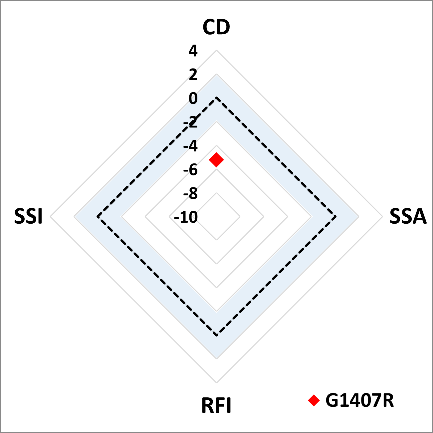

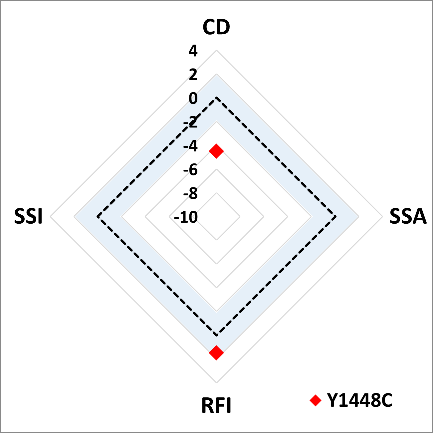

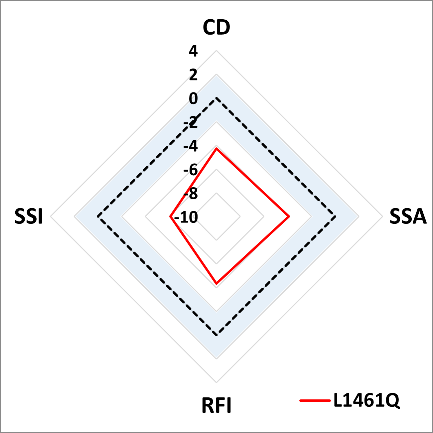

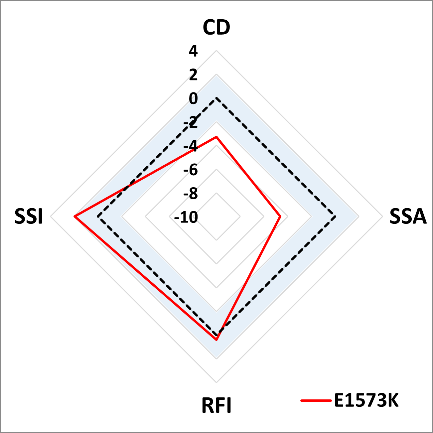

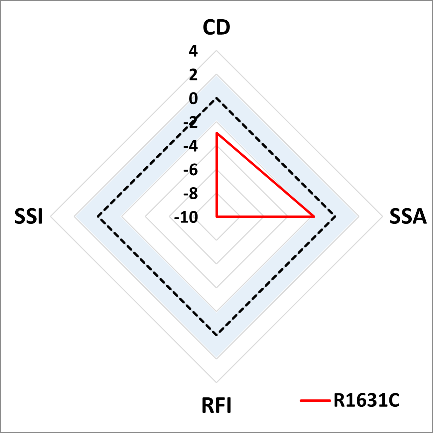

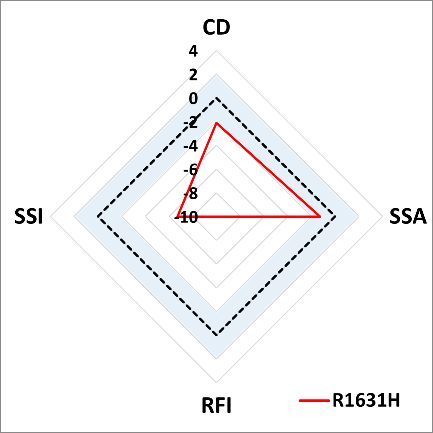

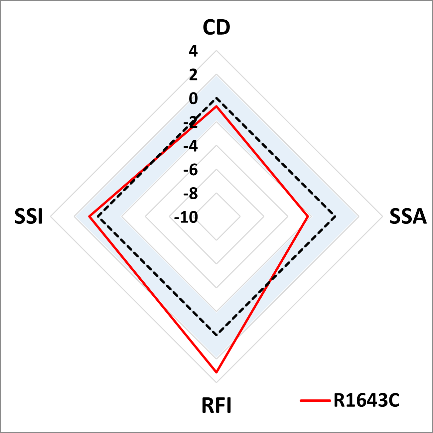

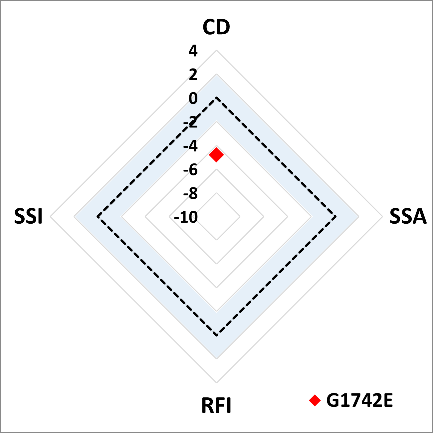

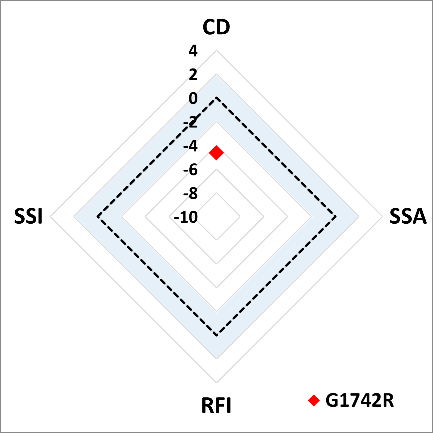

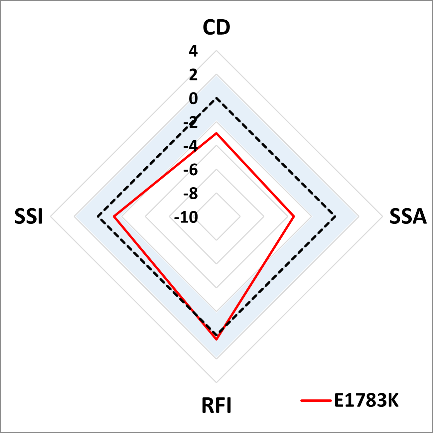

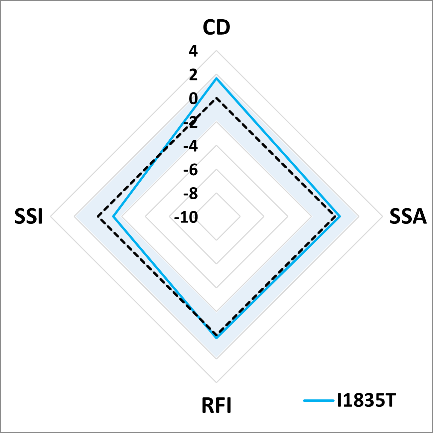

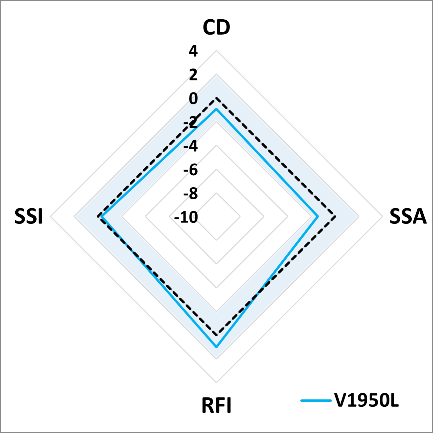

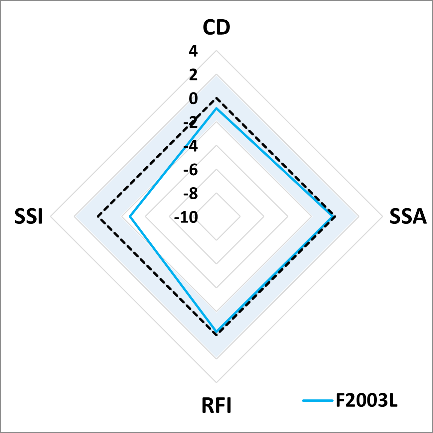

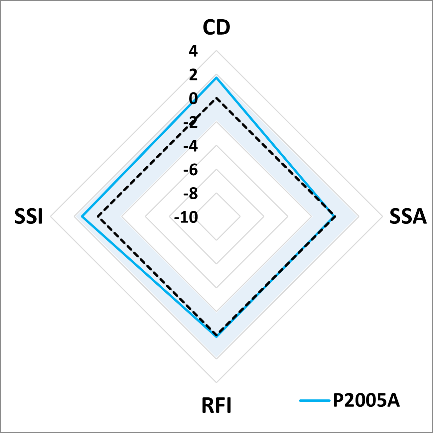
